# Prognostic whole-genome and transcriptome signatures in colorectal cancers

**DOI:** 10.1101/2023.03.28.23287846

**Authors:** Luís Nunes, Fuqiang Li, Meizhen Wu, Tian Luo, Klara Hammarström, Emma Lundin, Ingrid Ljuslinder, Artur Mezheyeuski, Per-Henrik Edqvist, Anna Löfgren-Burström, Carl Zingmark, Sofia Edin, Chatarina Larsson, Lucy Mathot, Erik Osterman, Emerik Osterlund, Viktor Ljungström, Inês Neves, Nicole Yacoub, Unnur Guðnadóttir, Helgi Birgisson, Malin Enblad, Fredrik Ponten, Richard Palmqvist, Mathias Uhlén, Kui Wu, Bengt Glimelius, Cong Lin, Tobias Sjöblom

## Abstract

Colorectal cancer (CRC) is caused by a sequence of somatic genomic alterations affecting driver genes in core cancer pathways^1^. To understand the functional and prognostic impact of cancer-causing somatic mutations, we analysed the whole genomes and transcriptomes of 1,063 primary CRCs in a population-based cohort with long-term follow-up. From the 96 mutated driver genes, 9 were novel to CRC and 24 to any cancer. Two distinct patterns of pathway co-mutations were observed, timing analyses identified 6 early and 3 late driver gene mutations, and several new signatures of CRC specific mutational processes were uncovered. Mutations in 10 protein-coding genes belonging to the WNT, EGFR, and TGF-β pathways, 2 mitochondrial DNA genes and 3 regulatory elements along with the COSMIC SBS44 signature impacted survival. Gene expression classification yielded 5 prognostic subtypes with distinct molecular features, in part explained by underlying genomic alterations. Microsatellite instable tumours could be divided in two classes with different levels of hypoxia and infiltration of immune and stromal cells. This study constitutes the largest integrated genome and transcriptome analysis of CRC to date, and links mutations, gene expressions and patient outcomes. The identification of prognostic mutations and expression subtypes can guide future efforts to individualize CRC therapy.

## Main

Colorectal cancer (CRC) is the third most common and the second deadliest tumour type in both sexes, with 1,900,000 new cases and 900,000 deaths annually. About 20% of patients have metastatic disease already at diagnosis, and another 20% will develop metastases later^2^. From exome^3–7^ and, to a more limited extent, whole-genome^8–10^ sequencing, the mutational landscape of CRC is best characterized in coding regions, whereas non-coding regions remain understudied. Approximately 80-85% of CRCs are classified as copy-number altered microsatellite stable (MSS), 10-16% as highly mutated tumours with microsatellite instability (MSI), and 1-2% as ultra-mutated tumours resulting from somatic *POLE* mutations. The MSI status predicts response to checkpoint inhibitors^11^, whereas *KRAS*, *NRAS* and *BRAF* mutations predict poor response to EGFR-targeted therapies^12^. The WNT, EGFR/KRAS/BRAF, PIK3CA, TGF-β, and P53 pathways are among those regulated by mutations in CRC^1^, and several driver gene mutations in these pathways have been linked to prognosis. The state-of-the-art gene expression classifier, Consensus Molecular Subtypes (CMS), divides CRCs into four subtypes which correlate with prognosis^13, 14^. To advance the understanding of CRC pathogenesis, identify driver events, and find prognostic features, we analysed whole genomes along with tumour transcriptomes in a large, population-based CRC cohort with clinical outcomes.

### Mutational landscape

We obtained high quality whole-genome sequences (average 53-fold coverage) from patient-matched tumour and normal samples from 1,063 CRC cases, along with tumour transcriptome sequence (average 30 M paired reads; **Table 1** and **Supplementary Table 1**). Of these, 943 were primary tumour surgical specimens and 120 were primary tumour biopsies. Of all patients, 126 (12%) had been pre-treated before the tumour specimens were obtained at surgery, and 92 of these (73% of the pre-treated) were rectal cancers treated with either chemoradiotherapy or radiotherapy prior to surgery.

**Table 1.** Patient and tumour characteristics by tumour hypermutation status.

| **Characteristics** | **All subjects**<br>**n = 1,063 (100%)** | **Non-Hypermutated**<br>**n = 821 (77%)** | **Hypermutated**<br>**n = 242 (23%)** | **P-value<sup>#</sup>** |
| --- | --- | --- | --- | --- |
| **Age (years)** |  |  |  |  |
| Mean ± S.D. | 71.2 ± 11.3 | 70.3 ± 11.1 | 74.3 ± 11.3 |  |
| Median (Range) | 72 (28-94) | 71 (28-94) | 76 (30-94) |  |
| >75 years | 422 (40%) | 293 (36%) | 129 (53%) | **8.65e-6** |
| **Sex** |  |  |  |  |
| Female | 514 (48%) | 356 (43%) | 158 (65%) | **2.35e-9** |
| **Primary Tumour Location** |  |  |  |  |
| Right Colon | 498 (47%) | 301 (37%) | 197 (81%) | **<2.2e-16** |
| Left Colon | 284 (27%) | 249 (30%) | 35 (15%) |  |
| Rectum | 281 (26%) | 271 (33%) | 10 (4%) |  |
| **Histology Subtype** |  |  |  |  |
| Adenocarcinoma | 877 (83%) | 721 (88%) | 156 (64%) | **3.39e-15** |
| Mucinous adenocarcinoma | 186 (17%) | 100 (12%) | 86 (36%) |  |
| **Tumour Grade** |  |  |  |  |
| Low grade | 837 (79%) | 700 (85%) | 137 (57%) | **<2.2e-16** |
| **Tumour Stage** |  |  |  |  |
| Stage I | 138 (13%) | 106 (13%) | 32 (13%) | **1.09e-9** |
| Stage II | 392 (37%) | 261 (32%) | 131 (54%) |  |
| Stage III | 419 (39%) | 354 (43%) | 65 (27%) |  |
| Stage IV | 114 (11%) | 100 (12%) | 14 (6%) |  |
| **Mismatch Repair Status** |  |  |  |  |
| MSI | 223 (21%) | 2 (0%) | 221 (91%) | **<2.2e-16** |
| MSS | 840 (79%) | 819 (100%) | 21 (9%) |  |
| **Tumour Resection Surgery** |  |  |  |  |
| Yes | 1034 (97%) | 799 (97%) | 240 (99%) | 0.136 |
| **Pre-Treated** |  |  |  |  |
| Radiation | 76 (7%) | 73 (9%) | 3 (1%) | **6.53e-5** |
| Chemotherapy | 33 (3%) | 26 (3%) | 7 (3%) |  |
| Chemoradiotherapy | 17 (2%) | 13 (2%) | 4 (2%) |  |
| None | 938 (88%) | 709 (86%) | 228 (94%) |  |
| **Survival** |  |  |  |  |
| Median OS (95% CI) | 118 m (108-126) | 119 m (109-126) | 101 m (82-173) | 0.173 |
| 5-Year OS Rate Stages I-III % | 75 % | 74 % | 78 % |  |
| 5-Year OS Rate Stage IV % | 24 % | 24 % | 23 % |  |
| Median RFS (95% CI)* | 121 m (111-136) | 121 m (113-136) | 96 m (82-173) | 0.109 |
| 5-Year RFS Rate Stages I-III % | 69 % | 68 % | 74 % |  |

MSI, microsatellite instable; MSS, microsatellite stable; OS, overall survival; RFS, recurrence free survival; CI, confidence interval. \*RFS was calculated for stage I-III; <sup>#</sup>P-value from Fisher's exact test and log-rank test for survival comparisons between non-hypermutated and hypermutated cases, statistically significant ( $P < 0.05$ ) differences in bold.

In total, 96 mutated driver genes were identified, and 1,056 (99%) of the tumours had a somatic mutation in at least one of these (**Fig. 1a** and **Supplementary Table 2**). Based on total mutation count, 242 (23%) tumours were hypermutated (HM) with >23.16 mutations/Mb (**Fig. 1b** and **Supplementary Table 3**). The HM cases tended to be of higher age (median 76 vs 71 years) and females, and have right-sided, mucinous, high grade (poorly differentiated or undifferentiated cancer cells) and stage II tumours more often (**Table 1**)^15^. The MSI criteria (MSIsensor score ≥ 3.5) were fulfilled in 223 (21%) patients, of which only two were non-HM (nHM). In total, 15 HM tumours were MSS tumours with *POLE* or other mismatch repair gene mutations (**Fig. 1c**) and six were MSS tumours with high noncoding tumour mutation burden (TMB). Of the 96 driver genes, 65 were drivers in nHM tumours, 37 in HM, and 79 when all tumours were considered. From these, 19 driver genes were common to HM and the entire cohort, compared to 34 for nHM and entire cohort. The HM and nHM tumours shared 16 known CRC drivers (**Supplementary Table 2**).

**Figure 1.**
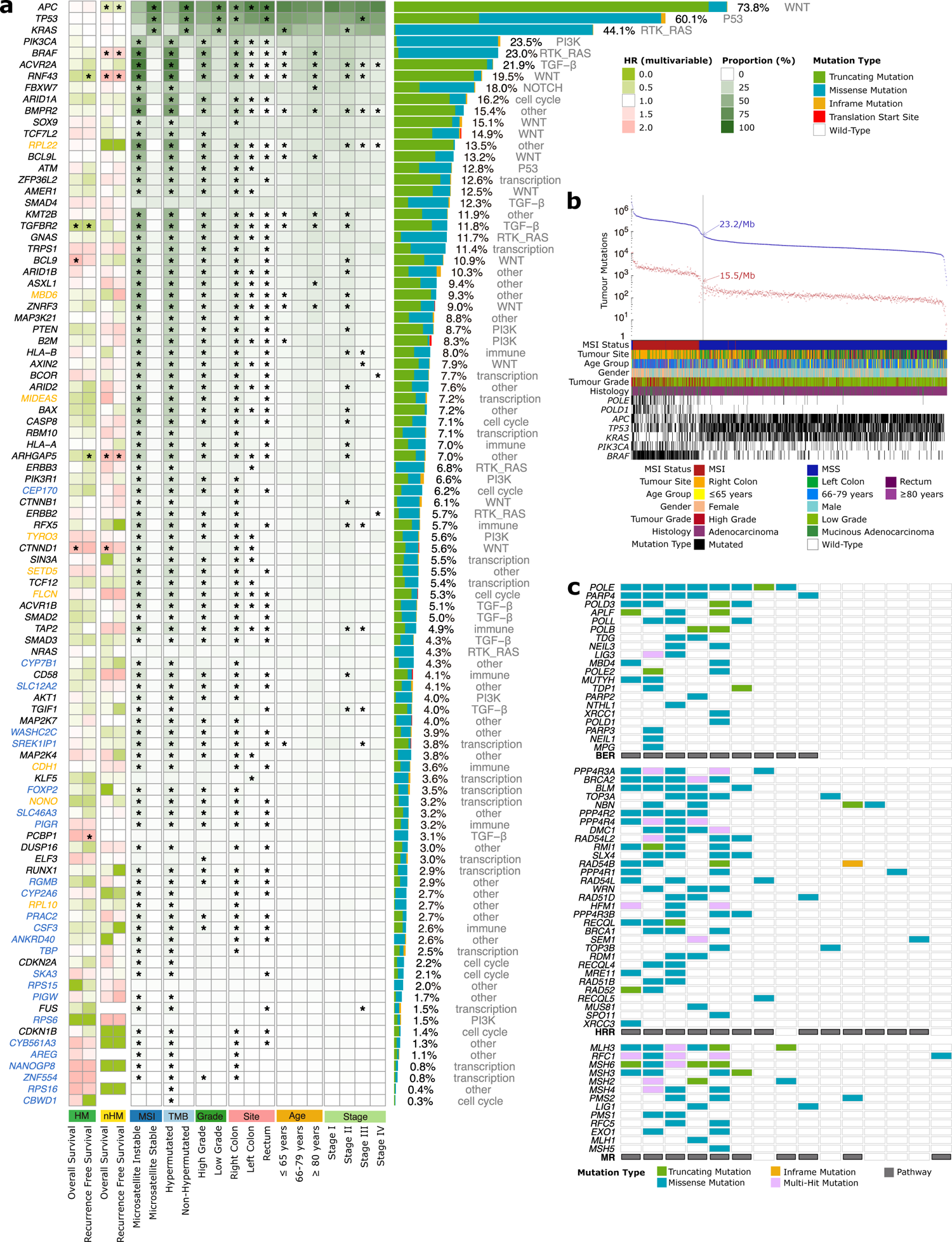
Somatic mutation analysis of 1,063 colorectal cancer genomes identifies 96 driver genes. Somatic mutations were called (see Methods) and significantly mutated genes identified using dNdScv. **a**, The 96 genes mutated at a significant level in this colorectal cancer (CRC) cohort. Association of driver genes with survival (HR) is shown for hypermutated and non-hypermutated tumours. The association of driver genes to clinical and genomic features is shown by proportion of tumours affected (* *FDR* or *P*<0.05). The mutation type and prevalence is indicated on the right, including a description of the affected pathway. Colour keys for hazard ratio for overall and recurrence free survival, and for genomic feature proportions are shown on the far right. Genes not previously designated as CRC drivers (orange) or not designated as driver genes in any cancer type (blue) are indicated. **b**, Prevalence of total (blue) and non-synonymous (red) mutations in each tumour. Cut-offs for hypermutated and non-hypermutated are indicated (grey line). Clinical features and mutation status for selected genes are shown at the bottom. **c**, DNA damage response gene mutations in the 15 of 21 hypermutated tumour cases that were microsatellite stable. HM, hypermutated; nHM, non-hypermutated; OS, overall survival; RFS, recurrence-free survival; HR, hazard ratio; MSI, microsatellite instability; MSS, microsatellite stable; BER, Base Excision Repair; HRR, homologous recombination repair; MR, mismatch repair.

In the HM tumours, genes mutated in >20% of the cases belonged to the TGF-β/BMP, WNT, RTK-RAS, ribosomal proteins, epigenetic regulation, PI3K, SCF complex, P53 and immune system pathways. Correspondingly, the WNT, P53, RTK-RAS, PI3K, SCF complex, and TGF-β pathways had genes mutated in >10% of nHM tumours (**Fig. 1a** and **Supplementary Table 2**). The most common hotspot mutations were *KRAS* G12D (15%) and G12V (11%) in nHM tumours, and *BRAF* V600E (65%) in HM tumours.

Of the 96 driver genes, the 24 that had not previously been designated as drivers in any cancer^16, 17^ were linked to BMP (*RGMB*) and EGFR (*AREG*) signalling, cell cycle (*CEP170* and *SKA3*), immune system (*PIGR* and *CSF3*), ion transport (*SLC12A2* and *CYB561A3*), metabolism (*PIGW*, *CYP2A6* and *CYP7B1*), mRNA splicing (*SREK1IP1*), protein transport (*WASHC2C* and *SLC46A3*), transcriptional regulation (*FOXP2*, *NANOGP8*, *TBP* and *ZNF554*), ribosomal proteins (*RPS15*, *RPS16* and *RPS6*), and other (*CBWD1*, *PRAC2* and *ANKRD40*). Nine drivers that had not previously been observed in CRC^3–5, 8, 10, 18–22^ were genes linked to the immune system (*CDH1*), histone modification (*SETD5*), transcription regulators (*MIDEAS* and *NONO*), PI3K signalling (*TYRO3*), cellular response (*FLCN*), ribosomal proteins (*RPL10* and *RPL22*) and UCH proteinase (*MBD6*).

Two distinct patterns of RTK/WNT pathway co-mutations, *KRAS*/*APC*/*AMER1* and *BRAF*/*RNF43*, were identified (**Supplementary Fig. 1a** and **Supplementary Table 4**). For the *KRAS*/*APC*/*AMER1* group, the nHM tumours had co-occurring *PIK3CA* (*FDR*=1.98e-5) and *TCF7L2* (*FDR*=4.57e-4) and mutually exclusive *TP53* (*FDR*=1.06e-7) and *NRAS* (*FDR*=1.76e-6) mutations. In the *BRAF*/*RNF43* tumours, co-occurring mutations were observed in *ACVR2A* (*FDR*=0.06) in HM tumours, and *AKT1* (*FDR*=0.03) and *TYRO3* (*FDR*=0.08) in the nHM tumours (**Supplementary Fig. 1b**). In the TGF-β pathway, co-occurring mutations were seen between *SMAD2* and *SMAD3* (*FDR*=1.03e-10) in nHM tumours, while *TGIF1* co-occurred with *PIK3CA* (*FDR*=0.09) in the HM cases. In addition, the HM tumours had mutually exclusive mutations in *B2M* and *HLA-A* (*FDR*=0.07)^23^ and co-occurring mutations of *KMT2B* with *CD58* (*FDR*=0.01) and *ERBB3* (*FDR*=0.09). In all, we identified 33 additional CRC drivers, adding details to the mutational landscape of HM CRCs, and identified novel co-mutation patterns within and across CRC pathways.

### Structural variants and relative timing of genomic alterations

To encompass all types of genomic events in the progression of CRC, we compiled copy number variants (CNVs) and structural variants (SVs)^24^. The most common chromosome arm aberrations were gains of 7p and 20q, detected in in almost 50% of tumours, and loss of heterozygosity (LOH) of 17p, 18p and 18q in >40% (**Fig. 2a**). Novel focal CNVs identified in nHM tumours included deletions of 15q24.3 (24.5%) harbouring *MIR3713*, 22q12.3 (23.5%), and 8p11.22 (18.6%) containing several ADAM family protease genes. The frequency of gene CNVs was higher in nHM compared to HM tumours, affecting an average of 55% vs 11% of the 96 driver genes. Driver genes most frequently affected by CNVs were *GNAS* and *ASXL1*, for which 82% and 81% of nHM tumours, respectively, had gains and/or amplifications, and *SMAD4* (79%), *SMAD2* (77%) and *TP53* (76%), which had deletions and LOH. In HM tumours, the antigen presenting genes *HLA-B* (26%), *HLA-A* (25%) and *TAP2* (24%) had highest LOH frequency, while *TRPS1* (26%), *ACVR1B* (22%), *CYP7B1* (22%), *MBD6* (22%) and *ERBB3* (22%) had highest frequency of CNV gains (**Supplementary Fig. 2** and **Supplementary Table 5**). Deletions were the most common SV, primarily in high grade (*FDR*=0.011) and less in stage I (*FDR*=0.021) tumours. Translocations were more common in older patients (*FDR*=0.004), HM (*FDR*=0.014), MSI (*FDR*=0.003) and high grade (*FDR*=0.003) tumours. In contrast, inversions and tandem duplications were less common in HM (*FDR*=4.69e-17 and 0.0091), MSI (*FDR*=2.75e-16 and 0.029) and right-sided tumours (*FDR*=6.45e-8 and 0.005; **Fig. 2b**). Half of the SVs affecting driver genes were deletions, most frequently affecting tumour suppressor genes including *RUNX1* (n=38), *PTEN* (n=33) and *SMAD3* (n=30). The most frequently affected DNA repair gene was *RAD51B* (n=33), often inactivated in breast and ovarian tumours (**Fig. 2c**)^25^. Extrachromosomal DNA (ecDNA) was observed in 250 tumours (24%), of which 91% were nHM (*P*=2.9e-9). Circular amplicons were observed in 87 (35%) of ecDNA+ cases, and the oncogenes most frequently harboured were *ERBB2* (n=9; 10%), *FLT3* (n=7; 8%), *CDX2* (n=7; 8%), *CDK12* (n=5; 6%) and *MYC* (n=5; 6%; **Supplementary Fig. 3a**). Tumour ecDNA conferred shorter survival in a pan-cancer study^26^, but no ecDNA-type dependent differences in overall survival (OS) or recurrence-free survival (RFS) were observed here (**Supplementary Fig. 3b**).

**Figure 2.**
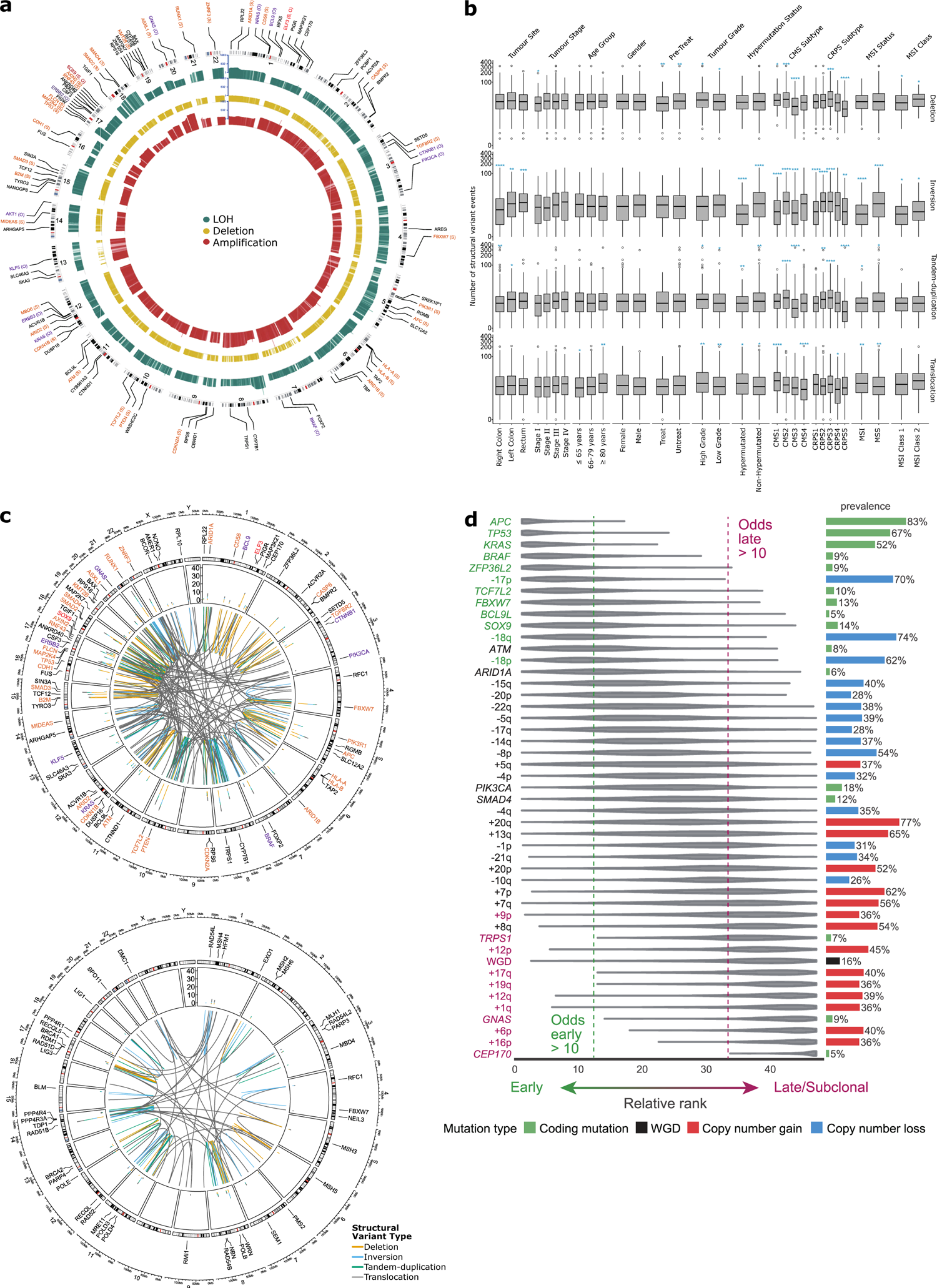
Structural variation and relative timing of somatic events in colorectal cancer. **a**, Gene copy number variants in driver genes displayed by type, loss of heterozygosity (green), deletion (yellow), and amplification (red). Bar height proportional to fraction of tumours with respective alteration. The 91 autosomal driver genes are indicated as oncogenes (purple), tumour suppressor genes (orange) or genes with unknown role (black) by genomic location. **b**, Structural variant (SV) landscape for deletions, inversions, tandem-duplications, and translocations by clinical, genomic and transcriptomic features (* *FDR*<0.05, ** *FDR*<0.01, *** *FDR*<0.001, **** *FDR*<0.0001, Wilcoxon test). **c**, Structural variants affecting driver genes (top) and DNA damage repair genes (bottom). Circos plots with counts (middle ring) for deletions (orange), inversions (blue), tandem-duplications (green) and translocations (yellow) by gene and chromosomal location. **d**, Prevalence and relative timing of driver gene mutations and SVs in 801 non-hypermutated CRC tumours by PhylogicNDT analysis. Early/clonal (green), intermediate (black) and late/subclonal (purple) alterations indicated. LOH, loss of heterozygosity; CMS, consensus molecular subtypes; CRPS, colorectal cancer prognostic subtypes; WGD, whole genome duplication.

The sequence of genomic events during CRC evolution has not previously been determined in a large set of carcinomas. We therefore investigated the relative timing of mutations and CNVs in nHM tumours^27^. The earliest events were somatic mutations in *APC*, *TP53*, *KRAS*, *BRAF* and *ZFP36L2*, followed by *TCF7L2, FBXW7*, *BCL9L* and *SOX9* and loss of chromosomes 17p and 18. Among them, (1) *TP53* and 17p loss are known early mutations frequently found in multiple cancers, (2) *APC*, *KRAS, BRAF* and *TP53* mutations drive CRC development, and (3) *ZFP36L2, TCF7L2, BCL9L* and *SOX9* are novel early events in cancer^28^. Late or subclonal events included whole genome duplication, gains of chromosomes 1q, 6p, 9p, 12, 16p, 17q and 19q, and mutations in *TRPS1*, *GNAS* and *CEP170* (**Fig. 2d**). These findings advance the understanding of CRC tumorigenesis and inform strategies for early detection and development of targeted therapies, and late events are of potential relevance for CRC invasion and metastasis.

### Mutational signatures

Mutational signatures in CRC have been linked to normal aging, mismatch repair (MMR) deficiency, polymerase proofreading, colibactin exposure and unknown aetiologies^29^. Here, a *de novo* mutational signature analysis identified 27 single-base substitution (SBS; **Supplementary Fig. 4**), 8 doublet-base substitution (DBS; **Supplementary Fig. 5**) and 11 small insertions and deletions (ID; **Supplementary Fig. 6**) signatures that were decomposed against COSMIC signatures^29^ (**Fig. 3a-c** and **Supplementary Table 6-7**). Of the 27 SBS signatures, 25 decomposed to 32 COSMIC SBS signatures (**Fig. 3a** and **Supplementary Table 8-9**). The most prevalent SBS signatures were the clock-like SBS5 and SBS1 (in >99% of the cases), followed by SBS18 (64%) which is linked to damage by reactive oxygen species (ROS). A new signature, SBS96J, was found in 17 tumours, all MSI or *POLE* mutants, and correlated with defective DNA MMR SBS15 signature (r=0.40, *FDR*=7.82e-40; **Supplementary Table 10**). Another, new signature SBS96AA, was observed in 17 cases with low grade and MSS tumours, and correlated with the UK_SBS128 signature of unknown aetiology (cosine similarity=0.94; **Supplementary Table 11**)^30^. Interestingly, HM tumours with the DNA MMR SBS44 signature were primarily located in the right colon (85% vs 70%, *P*=0.0064), less frequently stage IV (4% vs 15%, *P*=0.0386) with more *BRAF*-V600E (70% vs 45%, *P*=0.0015) and had longer OS (multivariable-HR=0.983, 95% CI: 0.967-1; **Fig. 3d**) than those without. Of the eight DBS signatures, three decomposed to six COSMIC DBS signatures while five could not be decomposed (**Fig. 3b** and **Supplementary Table 8-9**). The most prevalent DBS signature was the new DBS78A, present in 96% of all cases, while the new DBS78C had the highest somatic mutation density and occurred in 98% of the MSI cases. The defective DNA MMR signatures SBS15 and SBS44 strongly correlated with DBS78C (r=0.61 and 0.84, *FDR*=3.88e-108 and 8.11e-28) that was similar to the UK_DBS19 MMR deficiency signature (cosine similarity=0.87; **Supplementary Table 11**). The signatures SBS10a and SBS10b, associated with MSS *POLE* mutated tumours, strongly co-occurred with SBS28^31^ (r=0.48 and 0.61, *FDR*=5.13e-62 and 5.04e-107) and the novel DBS78G (r=0.43, *FDR*=8.21e-47; **Supplementary Table 10**). Lastly, from the eleven ID signatures, nine decomposed to nine COSMIC ID signatures, of which the most frequent, ID1 (in 87%) and ID2 (in 98%), are related to DNA slippage during replication^29^ (**Fig. 3c** and **Supplementary Table 8-9**). Interestingly, the novel ID83D signature had the highest somatic mutation density (>10 mutations/Mb), and 89% of the cases with ID83D also had the defective DNA MMR signature SBS44. Taken together, 47 known and nine novel (**Fig. 3e**) mutational signatures were identified, of which SBS28 and the new DBS78G signature were associated with *POLE* mutant MSS CRC, the SBS96J, DBS78C and ID83D signatures with MMR phenotype, and the DNA MMR SBS44 signature in HM tumours with longer OS.

**Figure 3.**
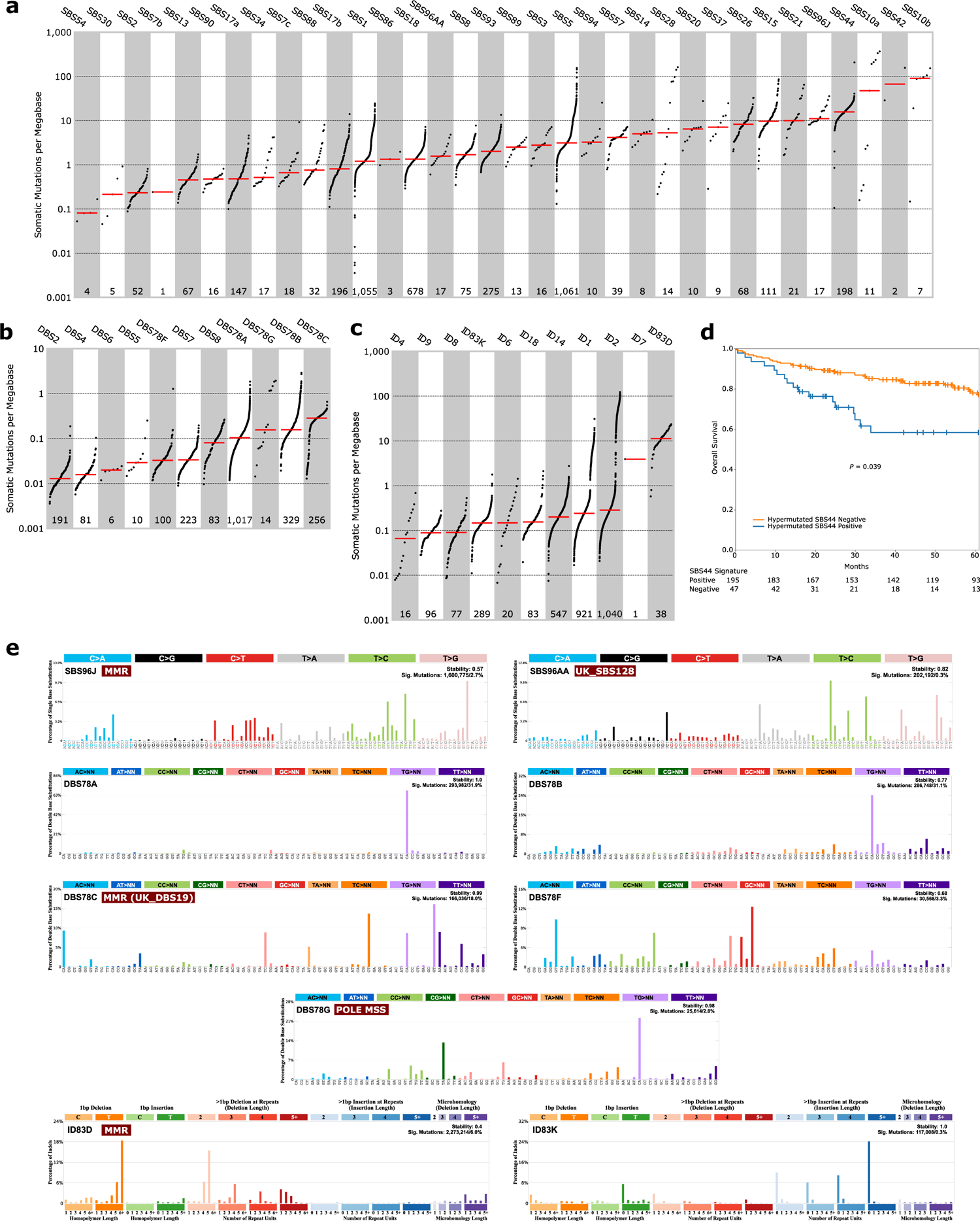
Identification of novel and prognostic somatic mutational signatures. *De novo* signature extraction and Cosmic signature decomposition by SigProfilerExtraction. Signatures of **(a)** single-base substitution, **(b)** doublet-base substitution, **(c)** and small insertion and deletion sorted by median (red line) mutational burden per megabase with each dot representing one tumour and the number of tumours with each signature indicated below. **d**, Overall survival of patients with stage I-IV hypermutated tumours (n=242) having the DNA mismatch repair SBS44 signature with Kaplan-Meier curves and log-rank test. **e,** SigProfilerExtraction profiles for the novel single-base substitutions (SBS96J and SBS96AA), doublet-base substitutions (DBS78A, DBS78B, DBS78C, DBS78F and DBS78G) and small insertions and deletions (ID83D and ID83K) signatures. SBS, single-base substitution; DBS, doublet-base substitution; ID, small insertions and deletions; MMR, mismatch repair; MSS, microsatellite stable.

### Mitochondrial genomes

High median copy number and enrichment of truncating mutations characterize CRC mitochondrial DNA (mtDNA) compared to other cancers^32^. We identified 3,982 single nucleotide variants (SNVs) and 949 indel mutations in mtDNA in 1,027 (97%) tumours (**Supplementary Table 12**). The mtDNA mutations were most frequent in the non-coding promoter D-loop (48%) and in the complex I genes *ND5* (41%) and *ND4* (31%; **Supplementary Fig. 7a**). Truncating mutations were enriched in *ND5* and *ND4,* representing 35% and 29% of their mutations respectively. Like in other cancer types^32^, missense mtDNA mutations were more frequently near homoplasmic (variant allele frequency, VAF >60%) compared with silent and truncating mutations (**Supplementary Fig. 7b**) and their overall d_N_/d_S_ ratio was close to 1 at different VAFs (**Supplementary Fig. 7c**). However, truncating mutations with VAF >60% occurred in 6.6% of tumours, compared to <3% in other cancers^32^, suggesting that mitochondrial dysfunction is important for CRC tumorigenesis. Similar to nuclear mutations, the mtDNA mutation load was correlated to age at diagnosis (**Supplementary Fig. 7d**). Co-occurrence but not mutual exclusivity was observed between mtDNA mutations (**Supplementary Fig. 7e**). Mitochondrial genome copy number (mtDNA-CN) was lower in right colon, high grade and hypermutated tumours (**Supplementary Fig. 8a**). When divided by low (n=127) and high (n=912) tumour mtDNA-CN (**Supplementary Fig. 8b**), there was a trend for longer OS in high mtDNA-CN cases but this was not statistically significant (**Supplementary Fig. 8c-d**). The mtDNA-CN was positively correlated with the clock-like (SBS1 and SBS5) and ROS (SBS18) SBS signatures but negatively correlated with most MMR signatures (**Supplementary Fig. 8e-g**).

### Prognostic alterations

Compared with cohorts from clinical trials, referral hospitals, or actively treated patients, patient age was higher (median 72 vs 54-68 years), right-sided tumours were more common (47% vs 30-39%) and the fraction of MSI cases was higher (21% vs 8-12%) in this cohort, leading to differential prognostic cohort features^4, 8, 18^. Rectal, stage I and stage IV tumours were slightly underrepresented but OS was similar when compared to all surgically resected CRCs in Sweden (**Supplementary Table 13**). In all, the cohort is representative of the resected Swedish CRC patient population, and as such of Western real-life populations. Within the cohort, the MSS tumour patients had shorter RFS (*P*=0.048) than MSI, while non-pre-treated stage IV MSI had shorter OS (*P*=0.039) than their MSS counterparts. The worst OS and RFS were observed in patients older than 80 years, as well as for high-grade and more advanced stage tumours. For non-pre-treated stage I-III patients, tumour location in rectum or left-sided colon correlated with longer OS (*P=*0.006) but not RFS (*P*=0.365). There was no survival difference by primary tumour location for stage IV CRC patients, although patients with right-sided tumours tended to do worse (**Supplementary Table 14**).

Since 768 (72%) patients, of which 158 (21%) were HM CRC, had at least 5-year survival data, we assessed the prognostic potential of somatic alterations. Several mutated driver genes showed statistically significant associations with OS and/or RFS (**Supplementary Table 15**). For both nHM and HM tumours, *CTNND1* mutations were linked to shorter OS (HR=2.55 and 1.96; 95% CIs: 1.29-5.04 and 1.03-3.73). In the nHM group, *APC* mutations conferred longer OS (HR=0.64, 95% CI: 0.47-0.87) and RFS (HR=0.68, 95% CI: 0.49-0.94)^33^, whereas *ARHGAP5*, *BRAF*^34^ and *RNF43*^35^ mutations were associated with shorter OS (HR=3.71, 1.59 and 2.37; 95% CIs: 1.51-9.16, 1.08-2.33 and 1.39-4.03) and RFS (HR=2.68, 1.65 and 2.03; 95% CI: 1.09-6.59, 1.08-2.53 and 1.07-3.85). In contrast, HM cases with *ARHGAP5* and *RNF43* mutations had longer RFS (HR=0.40 and 0.43; 95% CIs: 0.19-0.85 and 0.23-0.78). In the HM group, *TGFBR2* mutation conferred longer OS (HR=0.35, 95% CI: 0.20-0.63) and RFS (HR=0.34, 95% CI: 0.19-0.62), whereas *BCL9* conferred shorter OS (HR=2.16, 95% CI: 1.23-3.77), and *PCBP1* shorter RFS (HR=4.22, 95% CI: 1.40-12.74). Mitochondrial mutations also exerted prognostic effects, the nHM tumours with *MT-CYB* mutations had longer OS (HR=0.63, 95% CI: 0.42-0.96) and RFS (HR=0.66, 95% CI: 0.46-0.94; **Supplementary Table 16**), while mitochondrial *MT-CO3* mutations were independently associated with shorter OS (HR=2.42, 95% CI: 1.16-5.05) and RFS (HR=2.35, 95% CI: 1.17-4.70) in HM tumours but not any other mitochondrial mutation (**Supplementary Table 16**). We analysed candidate cis-regulatory elements (cCREs) in nHM tumours and identified seven proximal enhancer-like, one promoter-like, seven DNase-only and eleven CTCF-only elements along with 34 differentially expressed linked genes (**Supplementary Table 17**). Of the deregulated genes, *ID2* and *HS3ST1* had regulatory element mutations linked to shorter OS (HR=3.47 and 2.87, 95% CI: 1.60-7.51 and 1.26-6.57; **Supplementary Table 18**). Further, *DAPK1* was linked to an element with shorter RFS (HR=2.68, 95% CI: 1.25-5.73). In comparison, coding mutations were found in *ID2* in two cases, in *HS3ST1* in one case, and in *DAPK1* in 14 cases. Expression of *ID2* was previously found increased by WNT signalling after hypoxia induction, *ID2* targeting reduced CRC cell growth *in vivo*^36, 37^ and *DAPK1* loss was linked to invasiveness of CRC cells^40^, supporting their roles in CRC pathogenesis. Prognostic CNVs in nHM tumours included known events such as amplification of 20q11.1 and 20q11.21 and loss of 16p13.3 (**Supplementary Table 19**). Novel prognostic CNVs included amplification of 20q13.2 and losses of 4p16.1, 4q34.1, 4q35.1, 8p23.1, 11p15.5, 12q24.33, 17p12, 17p13.3, 17q21.31and 21p12. Amplifications conferred longer, and losses shorter, survival (**Supplementary Table 19**). Patients with nHM tumours with *SMAD4* SV deletion had shorter RFS (HR=2.13, 95% CI: 1.04-4.34), and those with *TCF7L2* SV translocations had shorter OS (HR=5.23, 95% CI: 2.26-12.09) and RFS (HR=7.50, 95% CI: 2.72-20.69; **Supplementary Table 20**). Altogether, we observed association with prognosis for 10 known cancer genes, 14 CNVs, 3 mitochondrial genes and 3 regulatory elements (see **Table 2** for a summary).

**Table 2.** Prognostic genomic features by hypermutation status.

| Feature | Non-Hypermutated |  | Hypermutated |  |
| --- | --- | --- | --- | --- |
|  | Longer survival | Shorter survival | Longer survival | Shorter survival |
| **Coding driver gene mutation** | *APC*<sup>OS, RFS</sup> | *CTNND1*<sup>OS</sup><br>*ARHGAP5*<sup>OS, RFS</sup><br>*BRAF*<sup>OS, RFS</sup><br>*RNF43*<sup>OS, RFS</sup> | *ARHGAP5*<sup>RFS</sup><br>*RNF43*<sup>RFS</sup><br>*TGFBR2*<sup>OS, RFS</sup> | *CTNND1*<sup>OS</sup><br>*BCL9*<sup>OS</sup><br>*PCBP1*<sup>RFS</sup> |
| **Mitochondrial gene mutation** | *MT-CYB*<sup>OS, RFS</sup> | - | - | *MT-CO3*<sup>OS, RFS</sup> |
| **Non-coding driver gene mutation** | - | *ID2*<sup>OS</sup><br>*HS3ST1*<sup>OS</sup><br>*DAPK1*<sup>RFS</sup> | - | - |
| **Copy number variation** | +20q11.1<sup>OS, RFS</sup><br>+20q11.21<sup>OS</sup><br>+20q13.12<sup>OS</sup> | -4p16.1<sup>OS</sup><br>-4q34.1<sup>OS</sup><br>-4q35.1<sup>OS</sup><br>-8p23.1<sup>OS</sup><br>-11p15.5<sup>RFS</sup><br>-12q24.33<sup>RFS</sup><br>-16p13.3<sup>RFS</sup><br>-17p12<sup>RFS</sup><br>-17p13.3<sup>OS, RFS</sup><br>-17q21.31<sup>OS</sup><br>-17q25.3<sup>OS, RFS</sup><br>-19p13.3<sup>OS</sup><br>-21p12<sup>OS</sup> | - | - |
| **Structural variant** | - | *SMAD4*<sup>OS</sup><br>*TCF7L2*<sup>OS, RFS</sup> | - | - |
| **Mutational signature** | - | - | SBS44<sup>OS</sup> | - |

OS, overall survival; RFS, recurrence free survival.

### Expression of driver genes and fusion genes

High quality genome and transcriptome sequences from the same large set of tumours enable integrated analyses of gene mutations and gene expression levels. Among the driver genes, the WNT pathway was enriched for prognostic genes, and *RNF43*, *AXIN2*, *SOX9*, *ZNRF3, CTNNB1* and *AMER1* had 55-334% higher expression in tumours while *TCF7L2*, *APC* and *CTNND1* had 15-24% higher expression in normal colorectal tissue (**Supplementary Table 21**). Tumours with *SOX9* or *TCF7L2* mutations had increased expression of the respective genes, while other mutant WNT pathway drivers had reduced expression (**Supplementary Fig. 9**). Mutant *AKT1*, *CEP170*, *CDKN2A*, *KRAS*, *NRAS*, *RFX5* and *TGIF1* tumours had higher expression of the respective genes, compared with wild-type tumours. Decreased expression coupled to the gene mutation in the tumour characterized several genes related to antigen presentation (*HLA-A*, *B2M* and *CDH1*), transcription regulation (*ASXL1* and *NONO*), apoptosis (*BAX*), histone modification (*KMT2B*) and ribosomal functions (*PPL22*) as well as *BRAF*, *BMPR2*, *PTEN*, *TGFBR2*, and *TP53* (**Supplementary Fig. 9** and **Supplementary Table 21**).

A total of 621 fusion transcripts were expressed in 338 nHM (41%) and 78 HM (32%) tumours, 17 of which were recurrent (**Supplementary Fig. 10a**). The most frequently fused genes were *PTPRK* (n=27 tumours), *RSPO3* (n=25), *SEPTIN14* (n=24), *FBXO25* (n=24) and *FBRSL1* (n=19; **Supplementary Fig. 10b**). Of the fusions, 15 were known CRC drivers including *NTRK*, *BRAF* and *ERBB2* fusions, *PTPRK*-*RSPO3* fusion, which was previously shown to promote tumour differentiation and loss of stemness (**Supplementary Fig. 10c**)^39, 40^, and the uncharacterized *FBXO25-SEPTIN14* fusion (**Supplementary Fig. 10d**). These data can support development of expression-based classifiers and circulating tumour DNA biomarkers based on driver mutations.

### Prognostic gene expression signature

Mutational and transcriptional data can be used to develop subtyping classifiers where the contributions of underlying genomic events are defined. The CMS is the state-of-the-art gene expression-based classification of CRC^13^, but as a majority of CRCs are composed of several CMS subtypes when deconvoluted, the prognostic and predictive power of CMS may suffer from its inability to fully capture tumour heterogeneity^41^. A redefined five-subtype classification at single-cell resolution, the iCMS, has been proposed where CMS4 tumours separate into two subgroups with different prognosis^7^. Since the CMS classification is based on 18 datasets generated using different technologies, we asked if unsupervised classification of the cases studied here would recapitulate the CMS or not. Unsupervised *de novo* classification produced a novel classifier of five distinct groups. In this classification, which we termed Colorectal Cancer Prognostic Subtypes (CRPS), 97% of CMS1 tumours were classified as CRPS1, CMS2 tumours were distributed between CRPS2 (39%) and CRPS3 (59%), 68% of CMS3 tumours were classified as CRPS5, and CMS4 tumours distributed between CRPS2 (38%) and CRPS4 (45%; **Fig. 4a** and **Supplementary Fig. 11**). Importantly, CRPS assigned all but 3 tumours, while 192 (18%) remained unclassified by CMS, most of which were assigned to CRPS1 and CRPS2. The CRPS1 group contained 88% of the HM cases, whereas only 56% of HM cases were classified as CMS1. Accordingly, CRPS1 tumours most often occurred in right colon (79%), in older (median 76 years) and female (61%) patients and had the highest prevalence of somatic SNVs and the lowest of CNVs (**Supplementary Table 1** and **Supplementary Fig. 12a-c**). The CRPS2 and CRPS3 subtypes were distributed equally between anatomical locations and had very low frequencies of *BRAF* mutations (**Supplementary Fig. 12a**). The CRPS4 tumours were often rectal (47%) and had stromal, TGF-β and WNT pathway activation (**Fig. 4b** and **Supplementary Table 22**).

**Figure 4.**
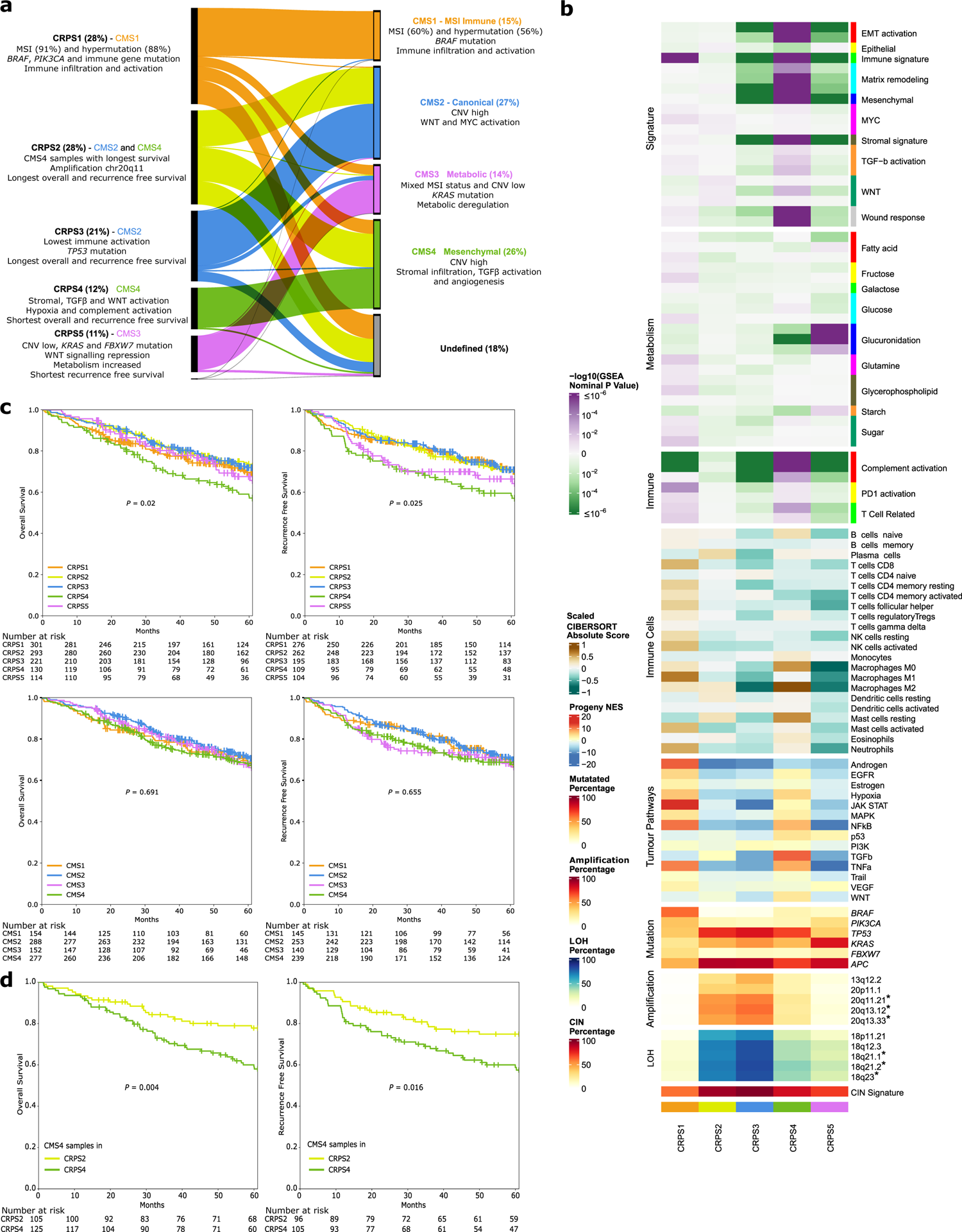
Refined prognostic subtypes derived from 1,063 colorectal cancer transcriptomes. The five distinct Colorectal Cancer Prognostic Subtypes (CRPS) yielded from unsupervised classification of tumour transcriptome data compared with classification of the same data set according to the Consensus Molecular Subtypes (CMS1-4). **a**, Comparison of CRPS to CMS. The proportion of samples assigned to each subtype is shown in percent. The main molecular and clinical characteristics for each CRPS and CMS subgroup are indicated. **b**, Transcriptomic characteristics of 1,063 samples according to their CRPS classification. Prognostic focal copy number variation cytobands that were differentially altered in CRPS are indicated by * (*P*<0.05, multivariable Cox regression in non-hypermutated samples). **c**, Kaplan-Meier survival curves (with multivariable log-rank test) and patients at risk for overall (stages I-IV) and recurrence free (stages I-III) survival in CRPS (top) and CMS (bottom) groups. **d**, Kaplan-Meier survival curves (with multivariable log-rank test) for CMS4 samples allocated to CRPS2 and CMS4 samples allocated to the CRPS4 group. MSI, microsatellite instability; CNV, copy number variation; LOH, loss of heterozygosity; CIN; chromosomal instability.

Finally, CRPS5 tumours were often from the right colon (46%), had WNT signalling repression (**Supplementary Table 22**) and the highest prevalence of *KRAS*, *PIK3CA* and *FBXW7* mutations, but fewer *TP53* mutations and CNVs compared with CRPS2-4 (**Fig. 4b** and **Supplementary Fig. 12a-b**). The distribution of re-clustered CRPS cases was robust to the removal of stage IV and pre-treated cases (**Supplementary Fig.13a**). The CRPS subtypes were prognostic regarding OS in stages I-IV (*P*=0.02) and for RFS in stages I-III (*P*=0.025; **Fig. 4c**). The CRPS2 and CRPS3 subtypes were associated with the longest OS and RFS, and CRPS4 with shortest OS and RFS, and CRPS5 the shorter RFS. The CMS4 cases assigned to CRPS2 had longer OS than those assigned to CRPS4 (**Fig. 4d**), which may reflect that the density of fibroblasts, macrophages and dendritic cells in CRPS2 is between those of CRPS3 and CRPS4 (**Fig. 4b**)^7^. For external validation, we developed a CRPS classification model based on ResNet50 (**Supplementary Fig. 14a**) and analysed gene expression data from 2,661 cases aggregated from 11 different cohorts. The accuracy, precision, recall and F1 score were >85% in the validation cases. Interestingly, 20q11 amplification was the top contributing feature for CRPS2 (**Supplementary Fig. 14b** and **Supplementary Table 20**)^42^. The prognostic ability of CRPS and the correspondence between CMS and CRPS was recapitulated (**Supplementary Fig. 13b-c**), with CRPS2 having the longest OS and CRPS4 the shortest (*P*=0.029 for all CRPS). Importantly, pathway features of CRPS subtypes were reproduced in the validation cohort (**Supplementary Fig. 13d**). Taken together, the CRPS outperforms CMS as gene expression classifier of CRCs, and can assign a very high proportion of tumours to subtypes.

### Tumour hypoxia

Among 27 tumour types previously evaluated, CRC ranked as the third most hypoxic^43^. To delineate links between genomic and transcriptomic alterations and tumour oxygenation, we analysed the transcriptomes using the Buffa hypoxia signature^44^. Tumours consistently had elevated hypoxia scores compared to normal CRC tissue (median 1 vs −20), where right colon tumours had the highest, followed by left colon and rectum (median 7 vs 1 vs −5; Fig. 5a). Consequently, tumours in females and of high grade, HM and MSI tumours were more hypoxic. Considering all tumours, 11 SBS, 5 ID and 2 DBS mutational signatures were linked to hypoxia. The strongest associations were SBS1, SBS5 and ID1, more prevalent in samples with low hypoxia (**Fig. 5b** and **Supplementary Table 23**)^43^. The MMR related signatures SBS44, SBS26, SBS14, SBS96J and DBS78C correlated with high hypoxia. In contrast, SBS18, related to damage by ROS, and ID18, related to colibactin exposure, were inversely correlated. The two ID mutational signatures associated with slippage errors during DNA replication were both correlated with hypoxia, with ID1 inversely correlated and ID2 correlated^43^. In nHM tumours, most driver genes, particularly *SMAD4*, *SMAD2* and *FBXW7,* correlated with high hypoxia. In HM tumours only *RGMB*, *SMAD2, AREG* and *RFX5* mutations correlated with low hypoxia (**Supplementary Fig. 15a** and **Supplementary Table 23**). Most SV types were associated with high hypoxia in nHM tumours^43^, but only deletions were associated with high hypoxia in HM tumours (**Fig. 5c** and **Supplementary Fig. 15b**). Elevated hypoxia scores were associated with higher number of clonal mutations in nHM samples, but not with the number of subclonal mutations (**Supplementary Fig. 15c**)^43^. High TMB was associated with hypoxia when considering all tumours. Impaired mitochondrial activity and abnormal mtDNA-CN characterized hypoxic tumours^45^, and mtDNA-CN was negatively correlated with hypoxia in both HM and nHM cases (**Supplementary Table 23**). Of the gene expression subgroups, CRPS1 and CRPS4 tumours were the most hypoxic, whereas CRPS3 and CRPS5 were the least. No correlation between hypoxia and patient survival was observed. In summary, these findings corroborate previous observations in nHM CRC and provide novel insights into hypoxia in HM CRC.

**Figure 5.**
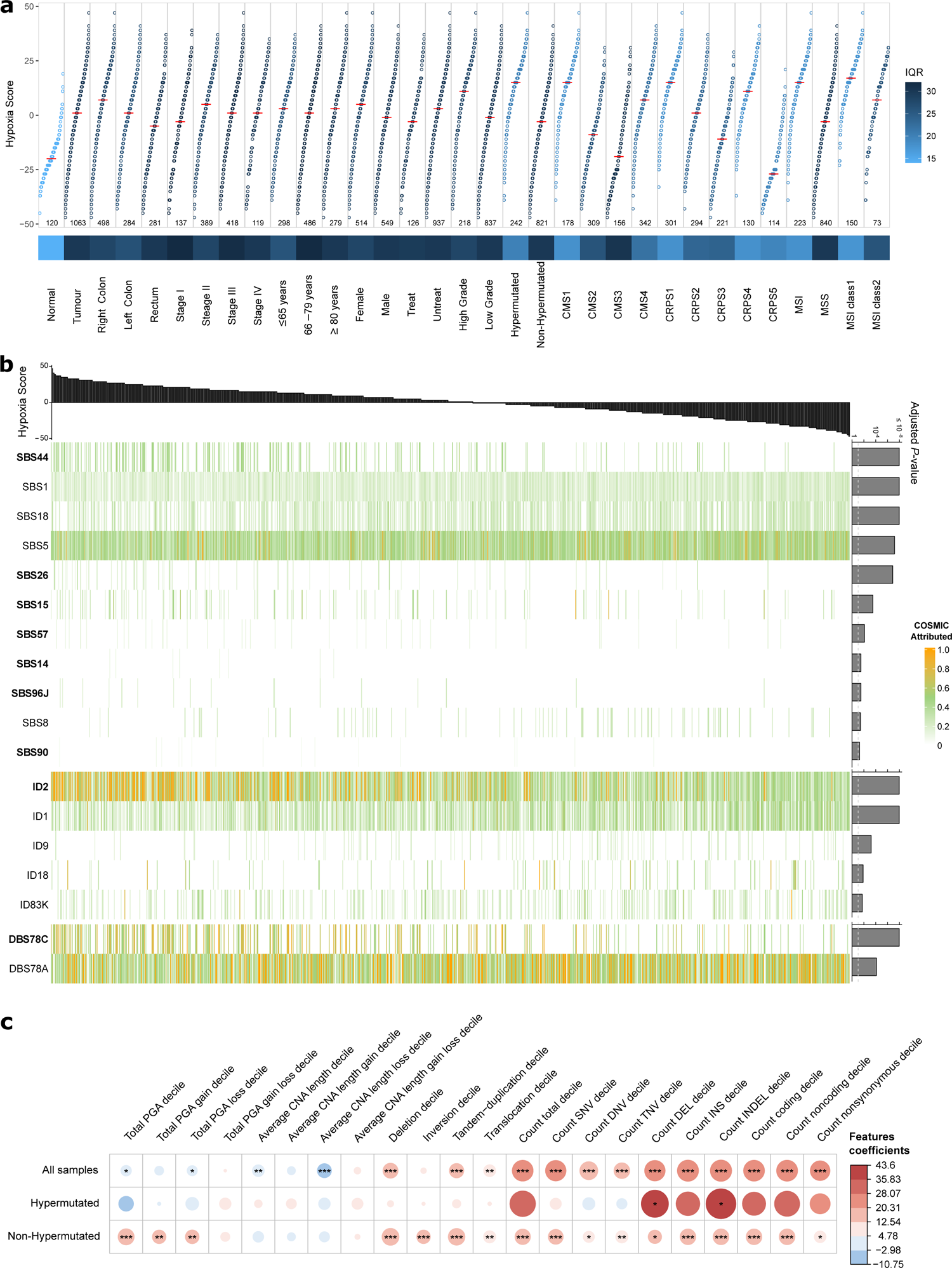
Hypoxia in colorectal cancer is associated with mismatch repair deficiency and genomic structural variation. **a**, Hypoxia scores based on the Buffa mRNA abundance signature for 1,063 tumour and 120 normal CRC tissues, by clinical, genomic and transcriptomic features. For each group, the median hypoxia score is marked (horizontal red line) and variability is coloured according to the interquartile range (IQR). **b**, Association of hypoxia score (top) with mutational signatures (bottom) coloured by normalized COSMIC signature activity attributed to each sample. Adjusted *FDR P*-values shown to the right and significance threshold indicated by dotted line. Signatures that showed positive correlation with the hypoxia score are shown in bold, the remainder displayed negative correlation with the score. **c**, Association of hypoxia scores with structural variants, by hypermutation status. Size and colour of the dots represent regression coefficients of the full model * *FDR*<0.05, ** *FDR*<0.01, *** *FDR*<0.001 (F-test full and null models comparison). IQR, interquartile range; PGA, percentage of genome with copy number alterations; CNA, copy number alterations; SNV, single nucleotide mutation; DNV, double nucleotide mutation; TNV, triple nucleotide mutation; DEL, deletion; INS, insertion; INDEL, insertion and deletion.

### Tumour microenvironment

The tumour microenvironment of each sample was characterised by transcriptome-based prediction of stromal and immune cell populations^46, 47^. The CRPS groups displayed differential infiltration of immune cells (**Supplementary Fig. 16a-b**). The CRPS1 tumours showed enrichment for T, B, dendritic cells and macrophages. Similarly, CRPS2 was characterised by elevated levels of hematopoietic stem cells (HSCs), dendritic cells and macrophages, while CRPS3 tumours had low levels of immune-cell infiltration but higher levels of megakaryocyte-erythroid progenitor cells (MEPs) and osteoblast-like cells. In CRPS4, there was an enrichment of fibroblasts, chondrocytes, endothelial cells, HSCs and macrophages, and low levels of epithelial, MEPs and T cells. The CRPS5 tumours were characterised by CD4 central memory and effector memory T cells. When stratified based on HM and MSI status, nHM tumours were characterised by infiltration of progenitor cells, such as hematopoietic stem and granulocyte-monocyte progenitor cells, and fibroblasts, but showed lower levels of mesenchymal stem cell and immune cell infiltration compared with HM cases (**Supplementary Table 24**). In nHM/MSS cases, the M2 like macrophages were associated with shorter OS and RFS, while T, dendritic and eosinophil cells were associated with longer OS and RFS (**Supplementary Table 25**).

In comparison to MSS tumours, MSI CRCs have high response rates to immunotherapy, but 45% of MSI tumours do not respond, motivating a finer grained subtyping^48^. We separated the MSI tumours into two classes using unsupervised classification (**Supplementary Fig. 17**), where the first was characterized by infiltrating lymphocytes and stromal cells, and the second by more abundant MEPs and T helper type 1 cell infiltration (**Supplementary Fig. 18**). The two MSI classes did not differ in OS or RFS, however, B cells were linked to shorter OS and RFS in MSI class 1 (multivariable HR>1), but not in class 2 tumours where M2 macrophages, CD4-T cells and erythrocytes were linked to shorter OS and RFS (multivariable HR>1; **Supplementary Fig. 19a** and **Supplementary Table 25**). MSI class 1 had more tumours carrying *ARID2* mutations, while class 2 contained more tumours with *BRAF* and SMAD4 CNV changes, *FOXP2* amplifications and 7q11 gains (**Supplementary Fig. 19b-d**). The MSI class 1 samples also had higher hypoxia scores (median 17 vs 7; **Fig. 5a**). These differences in immune cell composition and hypoxia levels motivate future analyses of immunotherapy responses in the two MSI classes.

## Discussion

This study is the largest that integrates WGS and transcriptome data from CRCs, all while providing sufficient clinical follow-up to enable analyses of prognostic factors. The molecular genetic basis of CRC is comparably well characterised, but the majority of analysed tumours stem from clinical trials, large referral hospitals or from tumours analysed prior to specific treatments. Here, we perform integrative analyses of CRCs that represent the incident patient population undergoing surgical removal of the primary tumour. Tumours with genetic alterations associated with poor prognosis are generally underrepresented in clinical trials where patient inclusion is based on specific criteria, as well as in hospital-based cohorts where only patients that may be eligible for treatment are analyzed^4, 8, 18^. We extend the CRC driver gene compendium by 33 genes, of which two-thirds were previously unknown to be cancer drivers, though, in the majority of instances belong to pathways of known importance in cancer. Several new mutational signatures related to defective DNA MMR and *POLE* mutations were identified. Timing analyses revealed that the vast majority of chromosomal losses are early events whereas amplifications are late events, and indicated that *TP53* mutation precedes *PIK3CA* mutation and loss of 10q (*PTEN*)^1^. Several novel early events were identified, and the late timing of amplifications and mutations in 1q and *CEP170*, 8q and *TRPS1*, and 20q and *GNAS* requires further analyses in the contexts of clonal fitness, invasion and metastases.

Analysis of multiple -omics modalities from the same tumours enabled deeper insight into prognostic features than either modality on its own. Important findings from the integrated analyses were that (i) the favourable CRPS2 and 3 type tumours were enriched for chromosome 20 amplifications previously linked to good prognosis^42^, (ii) M1 macrophages are enriched in the good prognosis CRPS1 type tumours and M2 macrophages in the poor prognosis CRPS4 type, (iii) key driver gene expression levels correlated with their mutation status, (iv) prognostic mutations in regulatory elements were linked to altered expression of specific genes, (v) tumour hypoxia was linked to specific mutational signatures and (vi) MSI tumours were divided into two classes with distinct molecular characteristics. Compared to current molecular classifiers, the prognostic CRPS signature provides a refined CRC subtyping into 5 classes with ability to classify the vast majority of tumours. Summarizing the prognostic genomic factors identified here (**Table 2**), we confirm the previously reported prognostic relevance of mutant *APC*, *BRAF* and *RNF43* and *SMAD4* loss in nHM CRCs^33–35, 49^, and report several novel prognostic driver genes, including the positive association with survival of the prevalent *TGFBR2* mutations in HM CRCs. Notably, the prognostic driver genes all belonged to the WNT, EGFR/KRAS/BRAF or TGF-β pathways. Further, the prognostic mtDNA mutations in *CO3* and *CYB*, and the mutations in the regulatory elements of *ID2*, *HS3ST1*, and *DAPK1* warrant further studies. Together, these findings provide fertile grounds for functional studies of CRC genes and for development of novel diagnostic and therapeutic modalities. Future characterisation of epigenomes, proteomes and metabolomes of the same tumours and patients can add additional insight into how different prognostic features are related to each other.

## Data Availability

Access to raw data and more detailed clinical information can be sought by contacting U-CAN (https://www.u-can.uu.se/?languageId=1). The remaining data are available within the Article, Supplementary Information or available from the authors upon request.

## Acknowledgments

We thank China National GeneBank (CNGB) and BGI-Henan for assistance with sequencing and computational resources. This study was funded by the Swedish Cancer Society (CAN 2018/772 and 21 1719 Pj to TS and 22 2054 Pj 01H to BG), the Uppsala Cancer Foundation to BG and by the Guangdong Provincial Key Laboratory of Human Disease Genomics (2020B1212070028). The U-CAN project, and part of this study, was funded by a grant from the Swedish Government (CancerUU) to Uppsala University, Umeå University, KTH Royal Institute of Technology, and Stockholm University (2010-ongoing), and grants from the Erling-Persson Foundation to TS and MU.

## Author contributions

L.N., F.L., K.H., P.E., A.M., V.L., H.B., M.E., F.P., M.U., B.G., C.L. and T.S. conceived the study; L.N., F.L., M.W., K.H., I.L., P.E., A.M., V.L., F.P., R.P., K.W., B.G., C.L. and T.S. coordinated the study; L.N., F.L., M.W., T.L., K.H., E.L., I.L., A.M., A.L., C.Z., S.E., L.M., E.O., E.O., I.N., N.Y., U.G. and C.L. performed research; L.N., F.L., M.W. and T.L. analysed data; L.N., F.L., T.L., K.H., P.E., C.L., F.P., R.P., K.W., B.G., C.L. and T.S. administrated and supervised the study; L.N., F.L., M.W., T.L., B.G., C.L. and T.S. wrote the paper with input from all other authors. All authors approved the manuscript before submission.

## Competing interests

Authors declare that they have no competing interests.

## Additional information

Supplementary Information is available for this paper. Correspondence and requests for materials should be addressed to.

## Notes

### Competing Interest Statement

The authors have declared no competing interest.

### Author Declarations

Sampling and analyses were performed under the ethical permits Uppsala EPN 2004-M281, 2010-198, 2007-116, 2012-224, 2015-419, 2018-490, and Umeå EPN 2016-219 and EPM 2019-566.

