## Supplementary material for "Prognostic whole-genome and transcriptome signatures in colorectal cancers": Methods

### *Patient cohort*

Patients diagnosed with CRC between 2004 and 2019, at the Uppsala University Hospital or the Umeå University Hospital, were eligible for the study. Patients that had i) a fresh frozen biopsy or surgical specimen that was estimated by a pathologist to have a tumour cell content of ≥20% and ii) a patient-matched source of normal DNA from whole blood or fresh frozen colorectal tissue stored in the biobank, were included. Clinical data was extracted from the national quality registry, the Swedish Colorectal Cancer Registry (SCRCR), and completed from medical records. Follow-up for alive patients was minimum 1 year and median 5 years (data lock 10$^{th}$ October 2020), with only one patient lost to follow up and 768 (72%) with complete 5-year follow up. Most patients included from June 2010 were drawn from the Uppsala-Umeå Comprehensive Cancer Consortium (U-CAN) biobank collections (Uppsala Biobank and Biobanken Norr)[1]. Sampling and analyses were performed under the ethical permits Uppsala EPN 2004-M281, 2010-198, 2007-116, 2012-224, 2015-419, 2018-490, and Umeå EPN 2016-219 and EPM 2019-566. Unfixed tissue materials from tumour and normal colon and rectum were handled on ice and frozen on the day of sampling or surgery[2]. Tissue pieces collected in Uppsala were embedded in Optimal Cutting Temperature (OCT) compound (Sakura, Japan) and stored at -70 °C. The tissue samples frozen in the Umeå University Hospital were frozen directly in pieces and stored at -70 °C, afterwards, embedded and cryo-sectioned for histology and tumour cell content confirmation. Haematoxylin-eosin (HE) stained sections from the frozen blocks were reviewed by a pathologist to confirm tumour histology and estimate tumour cell content. Patient-matched normal DNA samples were obtained from blood or frozen adjacent normal tissue. Normal RNA was obtained from 120 patient-matched colon or rectum tissue samples.

### *Tissue retrieval and nucleic acids extraction*

For Uppsala samples, five and eight cryosections of 10 µm each were used for RNA and DNA extraction, respectively. DNA was extracted using the NucleoSpin Tissue kit (cat. 740952; Macherey-Nagel, Germany), and RNA was extracted using the RNeasy Mini Kit (cat. 74106; Qiagen, Germany). For tissue samples from Umeå, DNA and RNA were extracted with AllPrep DNA/RNA/miRNA Universal kit (cat. 80224; Qiagen, Germany). Matching normal DNA samples were derived from peripheral blood (522 patients) or adjacent normal tissue (541 patients). Normal DNA from blood samples was extracted using the NucleoSpin 96 Blood Core kit (cat. 740456; Macherey-Nagel, Germany) on a Genomics STARlet robot (Hamilton, USA). For normal samples derived from tissue, DNA and RNA were extracted with the same procedures as described for the tumour samples. DNA concentration was measured using the Qubit broad-range dsDNA assay kit in the Qubit system (Invitrogen, USA), and RNA concentration and quality was assessed with Bioanalyzer RNA 6000 Nano kit (Agilent, USA) for samples from Uppsala and Tape Station 2200 (Agilent, USA) for samples from Umeå. RNA samples with RIN ≥7, 28s:18s ratio ≥0.8 and concentration ≥60 ng/µL were further analysed.

### *Whole-genome sequencing and data processing*

The WGS libraries were constructed from 1,063 primary CRC tumours and their paired normal samples according to the manufacturer's instructions for the MGIEasy FS DNA Library Prep Set (cat. 1000006987; MGI, China). The libraries were sequenced on a DIPSEQ platform (BGI, Shenzhen) and 100-bp paired-end sequencing was performed to yield data of ≥60× read coverage for all samples. During WGS data pre-processing, low-quality reads and adaptor sequences were removed by SOAPnuke (v2.0.7)[3] with parameters '-l 5 -q 0.5 -n 0.1 –f AAGTCGGAGGCCAAGCGGTCTTAGGAAGACAA -r

AAGTCGGATCGTAGCCATGTCGTTCTGTGAGCCAAGGAGTTG'. Sentieon Genomics software (version: sentieon-genomics-202010, https://www.sentieon.com/) was used to map and process high-quality reads for downstream analysis[4], which included the following optimised steps: i) BWA-MEM (version: 0.7.17-r1188) with parameters '-M -K 100000000' in alt-aware mapping model was used to align each tumour and normal sample to the human genome reference hg38 (containing all alternate contigs)[5]; ii) alignment reads were sorted by sort mode of Sentieon utility functions; iii) duplicate reads were marked by Picard (http://broadinstitute.github.io/picard/); iv) InDel realignment and base quality score recalibration for aligned reads were carried out by GATK[6]; v) and alignment QC was done by Picard.

***Somatic short variant calling***

Putative somatic SNVs, MNVs and/or INDELs were identified in each tumour-normal pair using multiple accelerated tools (TNhaplotyper, corresponding to MuTect2[7] of GATK3; TNhaplotyper2, corresponding to MuTect2[7] of GATK4; TNsnv, corresponding to MuTect[8]) and TNscope[9] of Sentieon Genomics software (version: sentieon-genomics-202010.01). Passed somatic SNVs, MNVs and INDELs detected by at least two tools were retrained as ensemble somatic short variants for each paired normal-tumour samples. Allele depths of ensemble somatic short variants were re-calculated by TNhaplotyper2 (version: sentieon-genomics-202010.01). High confidence ensemble somatic short variants (depth of tumour ≥14, depth of normal ≥8, variant allele reads count of tumour ≥2, variant allele reads count of normal ≤2, variant allele fraction of tumour ≥0.005 and variant allele fraction of normal ≤0.02) were selected for downstream annotation and analysis. These variants were annotated with VEP cache version 101 (corresponding to GENCODE v35) by Personal Cancer Genome Reporter (PCGR) (version: v0.9.1)[10].

***Somatic structural variants and copy number variation***

Somatic SVs were detected in each paired normal-tumour samples by BRASS (version: v6.3.4; https://github.com/cancerit/BRASS) with parameters '-j 4 –c 4 –s human –as GRCh38 –pr WGS', and ascatNgs[11] (version: v4.5; https://github.com/cancerit/ascatNgs) with parameters '-g L -q 20 -rs 'human' -ra GRCh38 -pr WGS -c 4 -force -nobigwig'. Genome cache file was generated by VAGrENT[12] (version: v3.7.0; https://github.com/cancerit/VAGrENT) with CCDS2Sequence.20180614.txt (https://ftp.ncbi.nlm.nih.gov/pub/CCDS/current_human/CCDS2Sequence.20180614.txt) and ensembl release-104 (http://ftp.ensembl.org/pub/release-104, Homo_sapiens.GRCh38.104.gff3.gz, Homo_sapiens.GRCh38.cdna.all.fa.gz, Homo_sapiens.GRCh38.ncrna.fa.gz). Other files for required parameters of BRASS and ascatNgs were extracted from CNV_SV_ref_GRCh38_hla_decoy_ebv_brass6+.tar.gz (ftp://ftp.sanger.ac.uk/pub/cancer/dockstore/human/GRCh38_hla_decoy_ebv/CNV_SV_ref_GRCh38_hla_decoy_ebv_brass6+.tar.gz). The SVs present in normal samples were filtered as follows. Somatic CNVs were detected in each paired normal-tumour sample by facetsSuite (version: v2.0.8; https://github.com/mskcc/facets-suite). An image of facetsSuite was pulled from docker://stevekm/facets-suite:2.0.8 and ran with singularity (v3.2.0)[13]. We used the aligned sequence BAM file as input data and executed FACETS in a two-pass mode with default settings in the R package[14]. First, the purity model estimated the overall segmented copy number profile, sample purity and ploidy. Subsequently, the dipLogR value inferred from diploid state in the purity model enabled the high-sensitivity model to detect more focal events. Allele specific copy numbers for each high confidence ensemble somatic short variants were annotated using the wrapper script 'annotate-maf-wrapper.R' with high-sensitivity output. Gene level copy number result was re-annotated with gencode v35.

Somatic copy number states were grouped into eight classes based on total copy number (tcn) and minor copy number (also known as lower copy number, lcn) estimated by FACETS, including wild type class (one copy per allele; tcn=2, lcn=1), homozygous deletions (tcn=0, lcn=0), loss of heterozygosity (LOH, tcn=1, lcn=0), copy-neutral LOH (tcn=2, lcn=0), gain-LOH (tcn =3 or 4, lcn=0), gain (tcn =3 or 4, lcn ≥1), amp-LOH (tcn ≥5, lcn =0) and amp (tcn ≥5, lcn ≥1).

### *Extrachromosomal DNA (ecDNA) detection*

Amplicons were detected in each sample by PrepareAA (commit ba747ce; https://github.com/jluebeck/PrepareAA) with parameters '--ref GRCh38 -t 4 --cngain 4.999999 --cnsize_min 50000 --downsample 10 --cnvkit_dir /home/programs/cnvkit.py --run_AA'[15,16]. Image of PrepareAA was pulled from docker://jluebeck/prepareaa:latest and ran with singularity (version: v3.2.0). The amplicons were then classified by AmpliconClassifier (version: v0.4.4; https://github.com/jluebeck/AmpliconClassifier) with parameters '--ref hg38 --plotstyle noplot --report_complexity --verbose_classification --annotate_cycles_file'[17]. The samples were classified based on which amplicons were present in the sample as previously described by Kim et al.[18].

### *Chromosomal instability signature quantification*

Activity of the 17 chromosomal instability (CIN) signatures presented by Drews et al.[19] were quantified by CINSignatureQuantification (version: v1.0.0; https://github.com/markowetzlab/CINSignatureQuantification) with unrounded copy number segments from facetsSuite. Tumours with normalised activities larger than zero, in any CIN signature, were identified as CIN samples.

***Microsatellite instability (MSI) detection***

The MSI status of CRC tumours was determined by running the MSIsensor2 tumour-normal paired module (v0.1, https://github.com/niu-lab/msisensor2) with parameters '-c 15 -b 4'. MSIsensor2 automatically detects somatic homopolymers and microsatellite changes and calculates MSI score as the percentage of MSI positive sites in all valid sites. Samples with MSI score ≥3.5 were considered MSI[20].

***Identification of significantly mutated genes***

Compared with other cancer types, hypermutated tumours associated with MSI or *POLE* mutation are frequently found in CRC. To avoid signals of samples with lower mutation burden from being masked during downstream WGS analyses, we first separated samples into hypermutated and non-hypermutated based on total count of somatic short variants according to the formula described previously[21]:

$$N_{SNV} > N_{median\_SNV} + 1.5 * IQR$$

After a first round of calculations based on the above formula, mutation counts in each detected hypermutated sample were split into two separate artificial samples with equal number of mutations. This process was iterated until no hypermutated samples were detected. Outlier times indicate how many times a sample was called as hypermutated in this process. The mutational heterogeneity caused by increased mutation burden of hypermutated tumours can reduce the power to detect driver genes and affect the identification of mutational signatures[22–24]. To identify CRC driver genes, we ran dNdScv[25] (commit dcbf8e5, https://github.com/im3sanger/dndscv) on the whole cohort and on hypermutated and non-hypermutated samples separately. A list of a priori known cancer genes (to be excluded from the indel background model) was constituted by COSMIC Cancer Gene Census[26] (v95) and intOGen Compendium Cancer Genes (Release date 2020.02.01, https://www.intogen.org/)[25,27–33]. Covariates (a matrix of covariates -columns- for each gene -rows-) were updated to covariates_hg19_hg38_epigenome_pcawg.rda (commit 9a59b89, https://github.com/im3sanger/dndscv_data). The reference database was updated to RefCDS_human_GRCh38_GencodeV18_recommended.rda (commit 9a59b89, https://github.com/im3sanger/dndscv_data). The dNdScv R package includes two different dN/dS-based algorithms, dNdSloc and dNdScv. The dNdSloc is like traditional dN/dS implementations, while dNdScv also takes into account variable mutation rates across genes and adds a negative binomial regression model using epigenomic covariates to infer the background mutation rate. The list of significant genes was selected by BH-adjusted *P*-values (qall_loc <0.1 or qglobal_cv <0.1) and merged from both dNdSloc and dNdScv. Long genes[34], olfactory receptor genes and genes with transcript per million (TPM) >1 in less than 10 samples were excluded from the potential driver gene list. Mutually exclusive or co-occurring sets of driver genes were detected using the modified somaticInteractions function of Maftools[35] (version: v2.12.0), which performs pair-wise Fisher's Exact test to detect significant (Benjamini-Hochberg False Discovery Rate (FDR) <0.1) pairs of genes.

***Identification of broad and focal somatic copy-number variation***

To determine significantly recurrent broad and focal somatic copy-number variants, GISTIC2.0[36] (v2.0.23) was run on resulting segmentation profiles from facetsSuite high-sensitivity models with parameters '-ta 0.3 -td 0.3 -qvt 0.25 -rx 0 -brlen 0.7 -conf 0.99 -js 4 -maxseg 25000 -genegistic 1 -broad 1 -twoside 1 -armpeel 1 -savegene 1 -gcm extreme -smallmem 1 -v 30'. A higher amplitude threshold according to GISTIC were used for focal copy number alterations classification, tumour and normal log2 ratio >0.9 for amplifications and <-0.3 for deletions[36]. Recurrently amplified or deleted regions were identified by GISTIC peaks and genes within each peak were summarized for further analyses.

### *Mutational signature analysis*

Analyses of mutational signatures were performed by SigProfilerExtraction[37] (version v1.1.4) with parameters '--reference_genome GRCh38 --opportunity_genome GRCh38 --minimum_signatures 1 --maximum_signatures 40 --nmf_replicates 500 --cpu 12 --gpu True --cosmic_version 3.2'. SigProfilerExtraction consists of two processes: *de novo* signature extraction and signature assignment[24,38,39]. Hierarchical *de novo* extraction of SBS, DBS, and ID signatures from all samples was followed by estimation of the optimal solution (number of signatures) based on the stability and accuracy of all 40 solutions. After signatures were identified, activities of each signature were estimated by assigning the number of mutations in each extracted mutational signature to each sample. SigProfilerExtraction also decomposed *de novo* signatures to the COSMIC[40] signature database[24] (version 3.2). The cosine similarity[41] between mutational signatures of the U-CAN and the GEL cohorts[42], and U-CAN and PCAWG cohorts[24] (COSMIC v3.3), were calculated with R (version v4.2.0). A *de novo* U-CAN signature was considered novel if the cosine similarity to both GEL and PCAWG signatures was <0.85. The mutational signature associations between U-CAN decomposed signatures were calculated by Stats::cor (method = "spearman") and corrplot::cor_mtest (conf.level = 0.95, "spearman") in R (version v4.2.0), and those with FDR $P$ <0.05 were considered statistically significant[43].

### *Analyses of non-coding somatic drivers in regulatory elements*

Regulatory elements were defined using SCREEN (Registry of cCREs V3, https://screen.encodeproject.org/), a registry of candidate cis-Regulatory Elements (cCREs) derived from ENCODE data[44]. Active cCREs annotated in 13 tissue samples (small intestine, transverse, sigmoid, left colon tissues) and 7 cell lines (CACO-2, HCT116, HT-29, LoVo, RKO, SW480 and HCEC 1CT) derived from colon were collected and downloaded from SCREEN, where cCREs are classified into six active groups (promoter-like signatures (PLS), proximal enhancer-like signatures (pELS), distal enhancer-like signatures (dELS), DNase-H3K4me3, CTCF-only and DNase-only) based on integrated DNase, H3K4me3, H3K27ac, and CTCF data. Further, the list of genes possibly linked to a cCRE according to experimental evidence (e.g., Hi-C) was extracted from the cCRE Details page of the website. Driver analyses were performed by ActiveDriverWGS[23,45] (commit 351ca77, https://github.com/reimandlab/ActiveDriverWGSR) with parameters '-mc 4 -rg hg38 -fh 300' on non-hypermutated samples for each cCREs groups. The missense mutations in the analyses of regulatory regions were removed to avoid confounding signals from known cancer drivers. Mutated elements with a Benjamini-Hochberg FDR <0.05 were considered to be significant and were used in the following analyses[45]. To evaluate the functional effects of driver cCREs, we examined their prognostic value and compared the expression levels of their linked genes. Cox proportional hazards analyses were performed to identify prognosis-associated cCREs using the Survival R package (version v3.3-1). Furthermore, potential associations between each cCRE and the expression levels of their linked genes were analysed by comparing raw expression values between groups of mutated and wild type samples using a two-sided Wilcoxon rank sum test. An FDR adjustment was applied to the *P*-values from the Wilcoxon test and genes with FDR <0.05 were considered to be differentially expressed with statistical significance. Finally, cCREs that had an impact on expression of linked genes were compared according to survival effects.

***Mitochondrial genome somatic mutation and copy number estimation***

We used multiple tools in GATK (version 4.2.0.0) workflow to extract the reads mapped to the mitochondrial genome from WGS, perform the mitochondrial DNA (mtDNA) variants calling and filter the output VCF file based on specific parameters, according to the official description (https://gatk.broadinstitute.org/hc/en-us/articles/4403870837275-Mitochondrial-short-variant-discovery-SNVs-Indels-). Further, false positive calls potentially caused by reads of mitochondrial DNA into the nuclear genome (NuMTs) were examined. These mutations normally have low variant allele frequency (VAF) but are highly recurrent in multiple tumours, as well as in matched normal samples. To remove these false positives, we employed stringent sample filtering, especially on variants with heteroplasmy <10%. We first performed two statistical tests as previously described[46]: i) the VAF of a mutation in the matched normal sequences needed to be <0.0034; and ii) the ratios of:

$$N_{mutnor}/RD_{nor} \,/\, (N_{mutnor}/RD_{nor}+N_{muttum}/RD_{tum})$$

needed to be <0.0629. These cut-offs were adapted from the same study and set by the median results of all mutation candidates plus 2 times the interquartile range. Since the occurrence rate of tumour-specific NuMTs is about 2.3%[47], we subsequently filtered the mutations whose frequency >0.023 in tumour samples. To avoid false negative calls in this step, the mutations with $VAF_{max}$ <0.1 and $VAF_{median}$ <0.05 were checked, and the samples in which the mutation had VAF >0.05 were kept[48]. The sequencing mean depth for the mitochondrial genome was 14,286x, allowing a better sensitivity for detection of somatic mutations at a very low heteroplasmy level, so the variants with 0.01<VAF<0.95 were used for the following analyses. For mtDNA copy number calculation, we used pysam (version 0.15.3) to filter and estimate the raw copy number of each sample. We then calculated the normalized copy number as previously described[49]. The survival best cut-point of mtDNA copy number was identified with surv_cutpoint (maxstat test: Maximally Selected Rank and Statistics) implemented in survminer (version 0.4.9). The associations between mutational signatures and mtDNA copy number were calculated by Stats::cor (method = "spearman") and corrplot::cor_mtest (conf.level = 0.95, "spearman") in R (version v4.2.0), and those with FDR $P$ <0.05 were considered statistically significant[43].

***Relative timing of somatic variants and copy number events***

For each non-hypermutated tumour, allele-specific copy-number-annotated high-confidence ensemble somatic short variants, and high-sensitivity copy-number events of autosomes (except the acrocentric chromosome arms 13p, 14p, 15p, 21p and 22p) were timed and related to one another with different probabilities using PhylogicNDT[50] (commit 84d3dd2, https://github.com/broadinstitute/PhylogicNDT). Single patient timing and the event timing in the cohort were inferred using PhylogicNDT LeagueModel as previously described[51]. The driver gene list identified in this cohort was specified to run PhylogicNDT.

***RNA-seq and determination of expression levels***

The rRNA was removed from total RNA using MGIEasy rRNA Depletion Kit (cat. 1000005953; MGI, China) and sequencing libraries were prepared for the 1,063 primary CRC tumours and 120 adjacent normal tissue samples with MGIEasy RNA Library Prep Kit V3.0 (cat. 1000006384; MGI, China) according to the manufacturer's instructions. Sequencing of 2 × 100 bp paired-end reads was performed using a DIPSEQ platform (BGI, Shenzhen) with a target depth of 30 M reads per sample. Pre-processing of RNA-seq data, including removal of low-quality reads and rRNA reads, was carried out using Bowtie2[52] and SOAPnuke. Clean sequencing data was mapped to human reference GRCh38 using STAR[53]. Expression levels of genes and transcripts were quantified using RNA-SeQC (version: v2.3.6)[54]. Transcripts with expression level 0 in all samples were excluded from further analyses and the mRNA expression matrix (19765*1183) was converted to $log_2$(TPM+1).

***Detection of oncogenic RNA fusions***

Gene fusions were detected by STAR-Fusion[55] (version v1.10.0; https://github.com/STAR-Fusion/STAR-Fusion) using clean FASTQ files with parameters '--FusionInspector validate --examine_coding_effect --denovo_reconstruct --CPU 8 --STAR_SortedByCoordinate' and Arriba[56] (version: v2.1.0; https://github.com/suhrig/arriba) starting with BAM files aligned by STAR[53] (version: v 2.7.8a; https://github.com/alexdobin/STAR). An image of STAR-Fusion was pulled from docker://trinityctat/starfusion:1.10.0 and ran with singularity (version: v3.2.0). Genome lib used in STAR-Fusion was downloaded from CTAT genome lib (https://data.broadinstitute.org/Trinity/CTAT_RESOURCE_LIB/__genome_libs_StarFv1.10/GRCh38_gencode_v37_CTAT_lib_Mar012021.plug-n-play.tar.gz). Aligned BAM files for Arriba were generated as described in the user manual (https://arriba.readthedocs.io/en/latest/). Gene fusions from Arriba were then annotated by FusionAnnotator (version v0.2.0, https://github.com/FusionAnnotator/FusionAnnotator) and merged with results of STAR-Fusion. Merged results were then filtered and prioritised with putative oncogenic fusions by annoFuse[57] (version v0.91.0; https://github.com/d3b-center/annoFuse).

***Unsupervised expression classification – CRPS generation***

We used Seurat (version 4.1.0) to identify stable clusters of all CRC samples, and among MSI tumours[58]. Potential batch effects or source differences between samples were corrected by Celligner[59] (https://github.com/broadinstitute/Celligner_ms), and the resulting matrix was imported into Seurat as scale data. Three different parameters were evaluated by repeating clustering with different k.param in FindNeighbors (10 to 30, step=5), number of principle components (10 to 100, step=5) and resolution in FindClusters (0.5 to 1.4, step=0.1). The stability of clusters was assessed by Jaccard similarity index and the preferred clustering result (resolution=0.9, PC=20, K=20) was determined by scclusteval[60] (version 0.0.0.9000).

### *Consensus molecular subtypes (CMS) classification*

For the CMS classification, three CMS classifier algorithms (CMSclassifier (version v1.0.0) with random forest prediction[61], CMSclassifier-single sample prediction[61], and CMScaller[62] (version v0.9.2)) were evaluated and results from the CMSclassifier-random forest was used. Expression data were processed using these three R packages separately or as combined, generating four sets of results. In the combined mode, the CMS subtype of each tumour sample was determined by at least 2 algorithms that predicted the same results, otherwise it was assigned as NA. Among all four sets of results, CMSclassifier-random forest predicted the most normal samples as NA and assigned more MSI samples to CMS1, indicating a lower false positive rate and a higher accuracy.

### *Model building and validation of CRPS classification*

To validate the CRPS *de novo* classification, we built a classification model based on the deep residual learning framework, involving the following steps. i) Gene expression data was first converted into pathway profiles by single-sample gene set enrichment analysis (ssGSEA[63]) implemented in Gene Set Variation Analysis (GSVA[64] (version v1.42.0), parameters 'min.sz=5, max.sz=300') using MSigDB[65–67] (version v7.4). We eventually obtained 30,049 pathways for 1,183 samples, including 1,063 tumours and 120 adjacent normal samples. ii) RelieF implemented in scikit-rebate[68] (version v0.62) was used to refine the obtained pathway features. The RelieF algorithm used nearest neighbour instances to calculate feature weights and assigned a score for the contribution of each feature to the CRPS classification. The features were then ranked by scores and the top 2,000 were selected for the model training. iii) We employed TensorFlow[69] (version v2.3.1) to construct the supervised machine learning model with a 50-layer residual network architecture (ResNet50), whose 4 stacked blocks were composed of 48 convolutional layers, 1 max pool and 1 average pool layer. During model compilation, we used the Nadam algorithm as the optimiser in terms of speed of model training and chose Categorical Crossentropy as loss of function in the classification task. In order to train the model sufficiently, epochs were set to 500 and LearningRateScheduler in Tensorflow was used to control the learning rate precisely in the beginning of each epoch; finally, ModelCheckpoint in Tensorflow was used to save the model with the maximum F1 score. iv) For the model training, the input data from 1,183 samples were divided into a training set (80%), a testing set (10%) and a validation set (10%). Batch size in Tensorflow was set to 6, corresponding to 5 clusters of CRPS and a normal sample cluster. To avoid bias caused by class imbalance during the learning process, Random OverSampling Examples algorithm in Imbalanced-learn[70] (version v0.9.0) was applied to ensure that at least one sample from each CRPS class could be randomly selected for model training. Samples with class probabilities less than 0.5 were categorised as NA. In addition, the Shapley Additive exPlanations (SHAP)[71] was applied to explain the model predictions on CRPS classifications, the molecular features of which could thus be interpreted. To test our CRPS clustering model, a total of 11 CRC data sets (n = 2,661 patients) from both NCBI GEO[72] (GSE2109, GSE13067, GSE13294, GSE14333, GSE17536, GSE20916, GSE33113, GSE35896, GSE39582 and GSE42284) and NCI Genomic Data Commons[73] (TCGA-COAD[22], TCGA-READ[22]) were uniformly processed from FPKM and transformed to pathway profiles with ssGSEA. After class prediction of these CRC samples by our CRPS clustering model, survival and pathway analyses were performed. Pathway analyses of CRPS from our dataset and from TCGA were performed with CMScaller[62]. The CRPS clustering model is available to use on https://github.com/SkymayBlue/U-CAN_CRPS_Model.

***Pathway analyses***

GSEA[65] (version v4.2.3 desktop) and MSigDB[66,67] (version v7.4) were used in pathway analyses, with the following settings: filter 'geneset min=15 max=200'. We also used PROGENy[74] (version 1.16.0) to investigate 14 oncogenic pathways in CRPS, as previously described.

***Hypoxia scoring and associations with mutational features***

Hypoxia scores were calculated for 1,063 CRC tumours and 120 normal samples, using the Buffa hypoxia signature[75] as previously described[76]. In brief, samples with mRNA abundance above the median tumour value of each gene in the signature were given a Buffa hypoxia score of +1, otherwise they were given a Buffa hypoxia score of -1. The sum of the score for every gene in the signature is the hypoxia score of the sample. We used a linear model to analyse the associations between hypoxia scores and mutational features of interest in all tumours, non-hypermutated tumours and hypermutated tumours using R stats package (version v4.1.0). For each mutational feature tested in the cohort, a full model and a null model were created and both were adjusted for tumour purity, age at diagnosis and sex[77]. The equations for the two models were adapted from the previous study[76] and shown below:

$$Full = hypoxia \sim feature + age + sex + purity$$

$$Null = hypoxia \sim age + sex + purity$$

Comparisons between the two models were made using ANOVA testing, and hypoxia was considered statistically significantly associated with a mutational feature when FDR or Bonferroni adjusted *P*-values were <0.1. Bonferroni adjustment was only applied to *P*-values when fewer than 20 tests were conducted. The scaled residuals for all full models were calculated by the simulateResiduals function in the DHARMa package[78] (version v0.4.5), and their uniform distributions were verified using the Kolmogorov-Smirnov test. Tested mutational features included mutational signatures, SNV, CNV and SV densities, driver mutations and subclonality. In the mutational signature analysis, the proportion of each signature in each tumour was used in the full model. To test the association between hypoxia and specific genetic alterations, we considered 22 metrics of mutational density in total, including: 10 SNV mutation counts of all regions, coding region, noncoding region, nonsynonymous, SNV, DNV, TNV, DEL, INS, and INDEL; 8 metrics of CNV mutational density which were adapted from the previous study by PCAWG[76], including the percentage of genome with total copy-number aberrations (PGA, total), PGA gain, PGA loss, PGA gain:loss, average CNV length, average CNV length gain, average CNV length loss and average CNV length gain:loss; and 4 SV types, including deletion, inversion, tandem-duplication, and translocation. Value of each decile for all 22 metrics were calculated with the R package dplyr[79]. Finally, in the subclonality analysis, clonal and subclonal mutations and numbers of subclones for each tumour were derived from PhylogicNDT as described above.

***Prediction of cell types in the tumour microenvironment***

The computational methods CIBERSORT[80] (version: v1.04) and xCell[81] (version: 1.1.0) were applied with default settings on TPM data for microenvironment estimation. The Intrinsic CMS (iCMS) subtype classification was performed as previously described[82]. In brief, 715 marker genes of intrinsic epithelial cancer signature were directly obtained from the previous research. The iCMS2 marker genes were taken from lists of iCMS2_up and iCMS3_down, and iCMS3_up and iCMS2_down lists were used as iCMS3 markers. Subsequently, scores of iCMS2 and iCMS3 for each tumour were calculated with 'ntp' function in CMScaller R package. Samples were defined as indeterminate if permutation-based FDR was ≥0.05.

***Survival analyses***

OS and RFS curves were constructed using the Kaplan-Meier method and the differences between groups were assessed by the log-rank test, using "survminer" package in R (version v0.4.9). OS was defined as time from diagnosis of primary tumour to death or censored if alive at last follow-up, while RFS as time from surgery to earliest local or distant recurrence date or death, or censored if no recurrence or death at last follow-up. The OS analyses included all stage I–IV patients, whereas patients with stage IV at diagnosis were excluded in the RFS analyses. Separate OS analyses were also performed for stage I-III for some variables. Cox's proportional hazards models were built to determine the prognostic impact of clinical and genomic features using "finalfit"/"survival" R packages (version v1.0.4/v3.3-1). Univariable Cox regression was performed on all identified coding or non-coding drivers and clinical variables, while multivariable Cox regression was applied on drivers that were statistically significant in the univariable analyses (*P* <0.05) with co-variates including tumour site, pre-treatment status, tumour stage, age groups, tumour grade, and hypermutation status. Forest plots were drawn using R "survivalAnalysis" (version v0.3.0) for visualising the prognostic value of tested features revealed by uni- or multivariable analyses. In the Supplementary tables showing associations with either OS or RFS, analyses showing *P*-values <0.05 were marked in bold. No compensation for multiple testing was done in these analyses.

***Data availability***

Access to raw data and more detailed clinical information can be sought by contacting U-CAN (https://www.u-can.uu.se/?languageId=1). The remaining data are available within the Article, Supplementary Information or available from the authors upon request. The patient identification numbers assigned to participants in this study were created solely for research purposes and are not known outside of this study. These IDs do not reveal the identity of the study subjects and are used solely for the purpose of data management and analysis.

***Code availability***

The CRPS clustering model is available to use on https://github.com/SkymayBlue/U-CAN_CRPS_Model.
