## Supplementary Figures for "Prognostic whole-genome and transcriptome signatures in colorectal cancers"

### Supplementary material

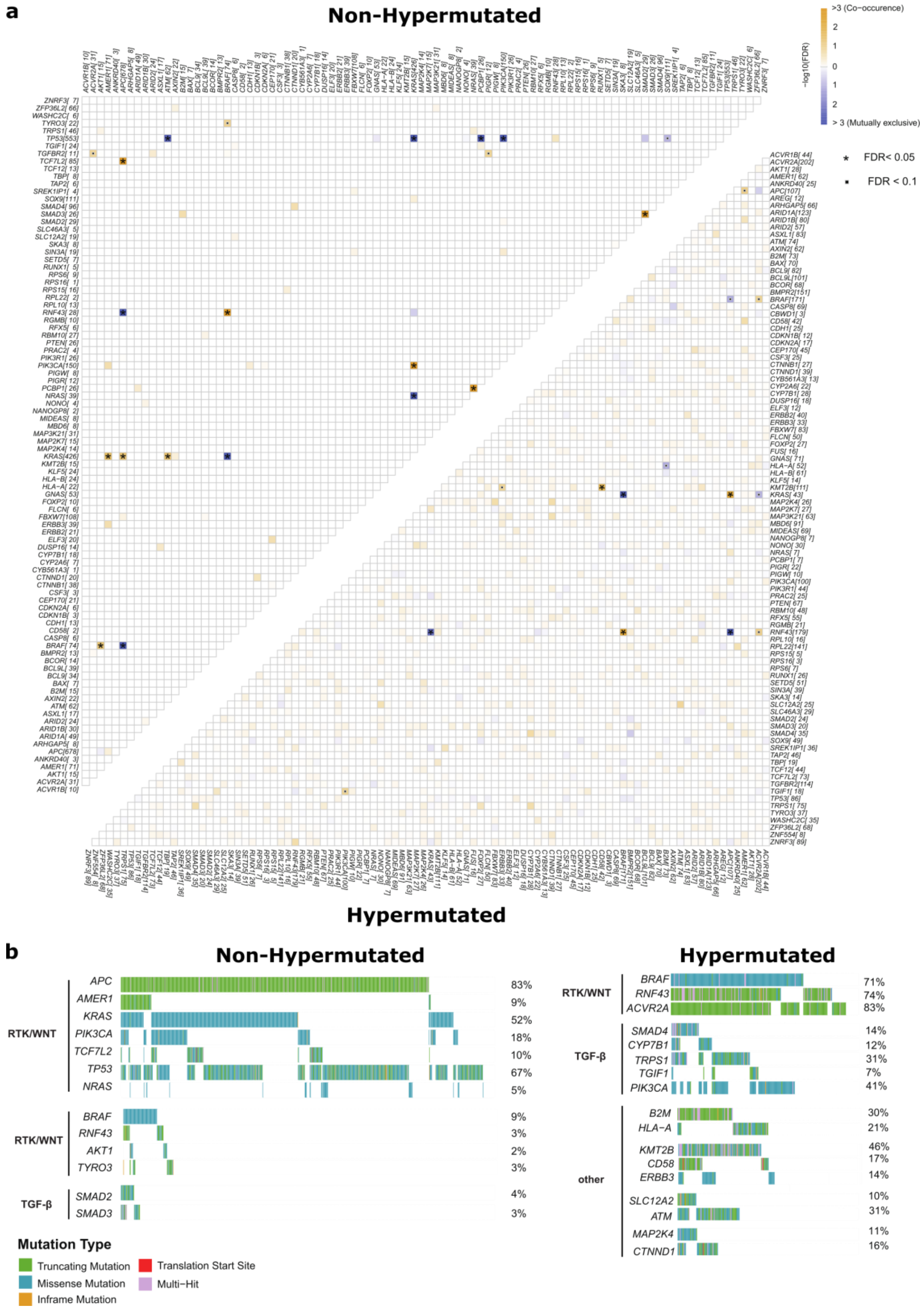


**Supplementary Figure 1. Mutually exclusive and co-occurring gene mutations in the 96 colorectal cancer driver genes by hypermutation status**.

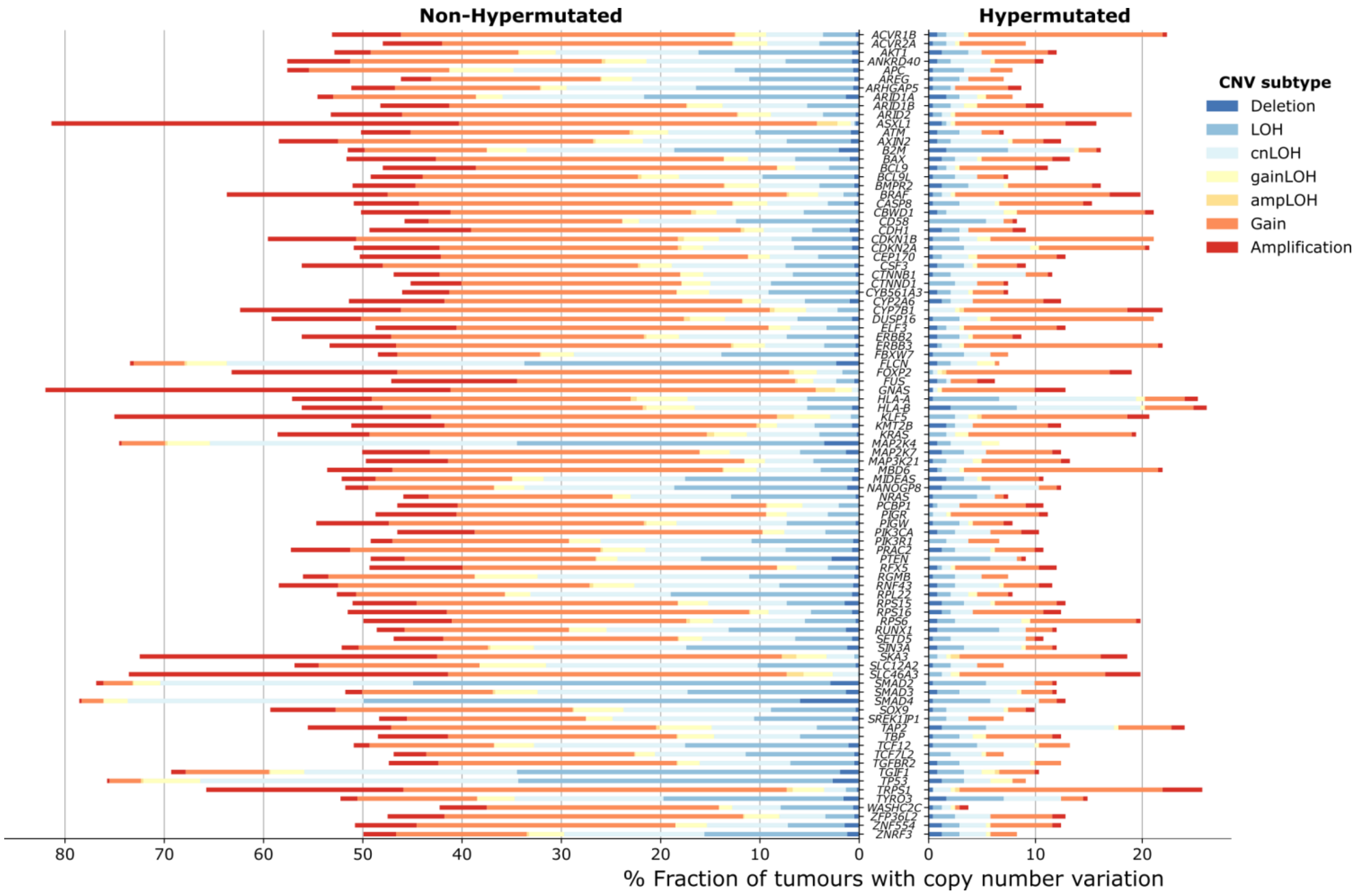


**Supplementary Figure 2. Copy number variation landscape for the 96 driver genes in non-hypermutated and hypermutated cases.** Copy number variation subtypes were called by facetsSuite. CNV, copy number variation; LOH, loss of heterozygosity; cnLOH, copy number neutral LOH; ampLOH, amplification LOH.

---

**Supplementary Figure 1 (continued). a**, Significant pairs of genes with mutually exclusive or co-occurring mutations were detected in non-hypermutated (n=821; upper half) and hypermutated (n=242; lower half) tumours with Fisher's Exact test adjusted by Benjamini-Hochberg False Discovery Rate (* FDR <0.05 and ▪ FDR <0.1). The number of patients with the mutation is represented inside brackets next to the gene name. **b**, Oncoplots displaying mutually exclusive and co-occurring driver gene mutations grouped by pathway in non-hypermutated (left) and hypermutated (right) tumours. Gene mutation prevalence is shown to the right.

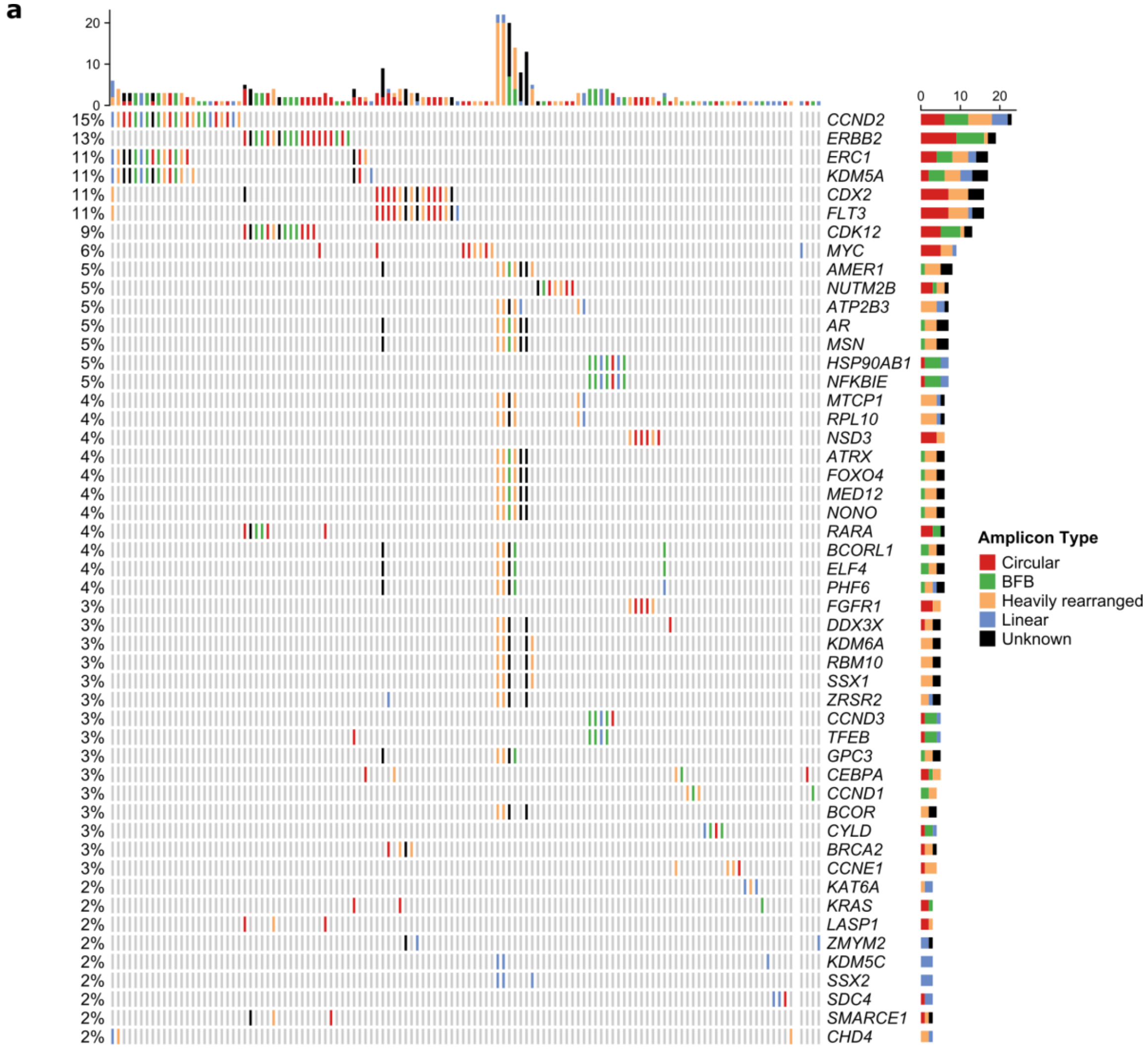


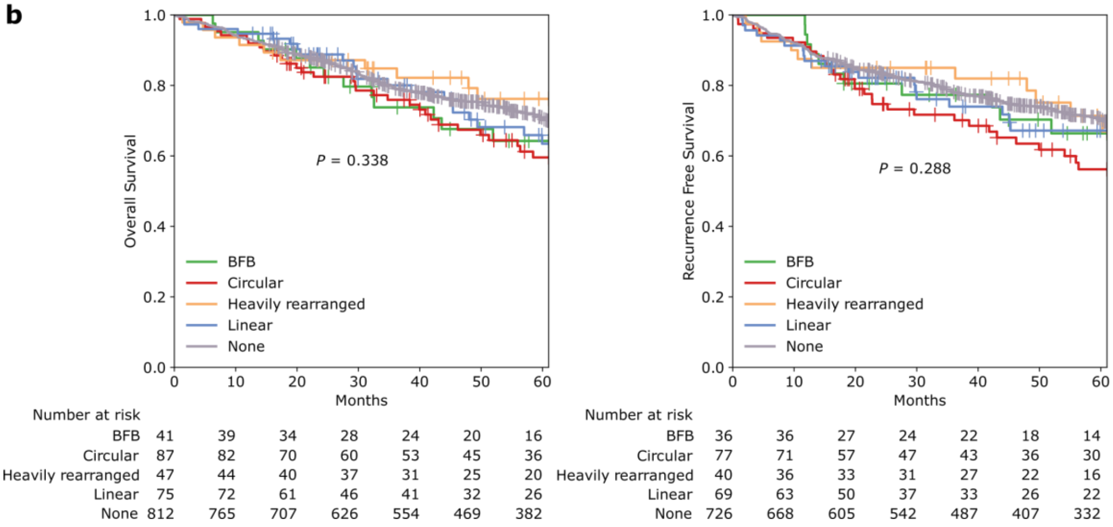


**Supplementary Figure 3. Extrachromosomal DNA landscape of colorectal cancer.** Tumours with extrachromosomal DNA (ecDNA) harbouring oncogenes were analysed. **a**, Oncoplot of the top 50 oncogenes observed with ecDNA amplicons in non-hypermutated (left columns) and hypermutated (last right 4 columns) tumours. Tot the left is shown the fraction of tumours with ecDNA involving the oncogene. Amount of ecDNA amplicons are represented by type for each tumour (top) and for each oncogene (right). **b**, Overall and recurrence free survival Kaplan-Meier curves for samples classified to be dominated by different types of ecDNAs. The tumours were classified based on which amplicons were present in the sample as previously described by Kim et al. (https://doi.org/10.1038/s41588-020-0678-2). Multivariate log-rank test with $P<0.05$ was considered significant. BFB, breakage-fusion-break.

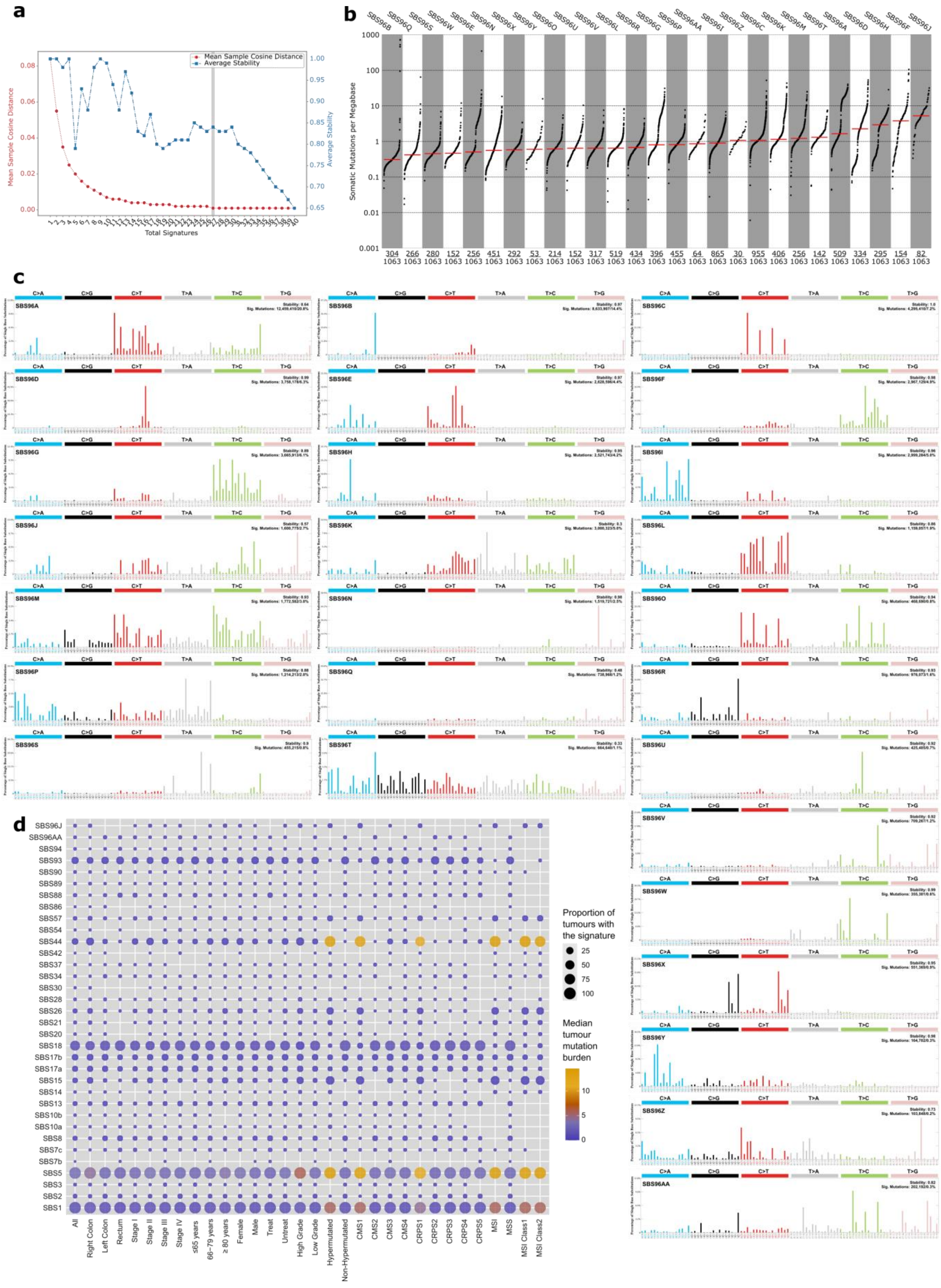


**Supplementary Figure 4. *De novo* extraction of single-base substitution (SBS) mutational signatures. a**, Hierarchical *de novo* extraction of SBS

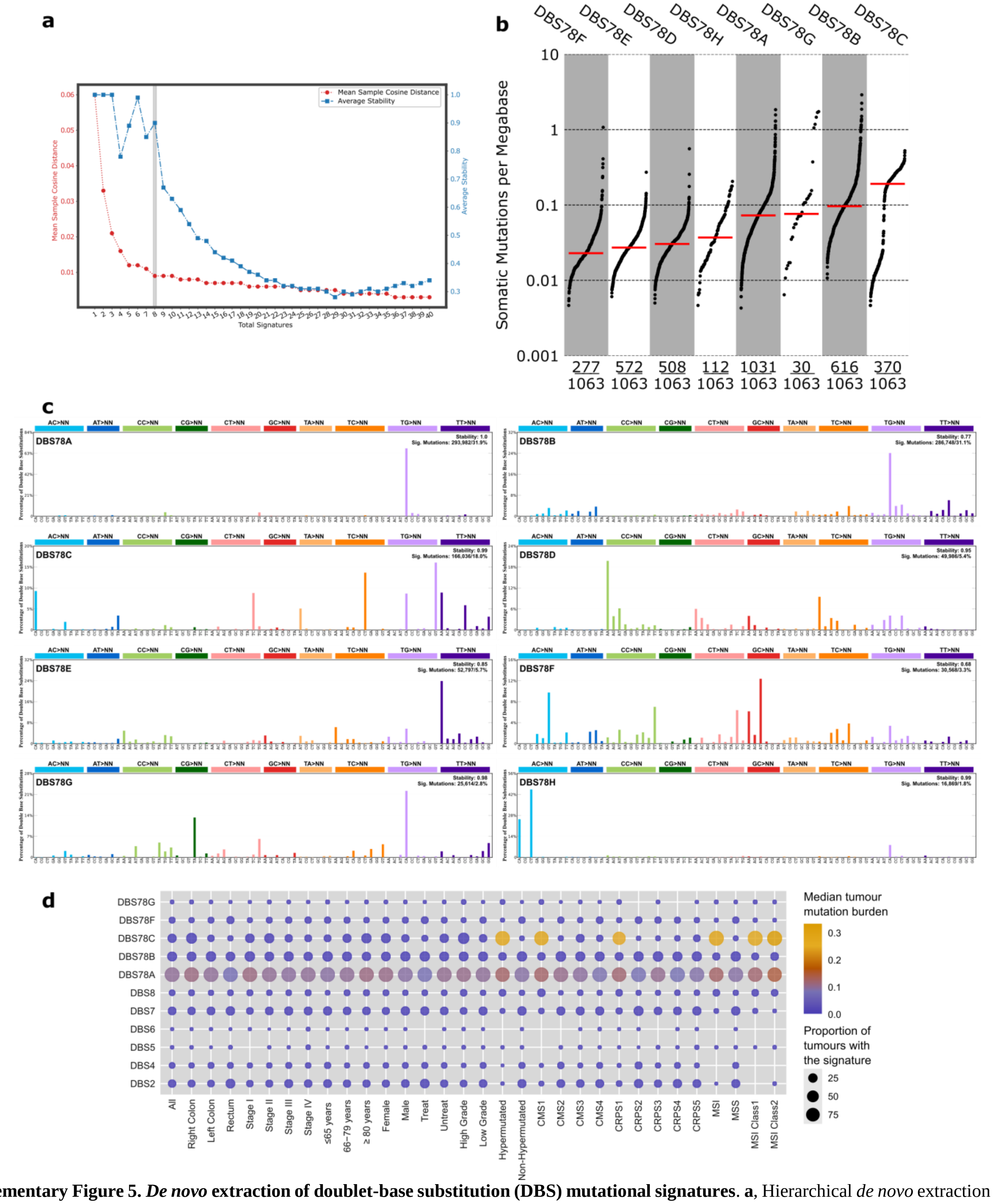


**Supplementary Figure 5.** ***De novo*** **extraction of doublet-base substitution (DBS) mutational signatures**. **a**, Hierarchical *de novo* extraction of DBS signatures from all tumours followed by estimation of the optimal solution (number of signatures marked with the grey line) based on the stability and accuracy of all 40 solutions. **b**, Mutation burden per megabase of each somatic DBS signature sorted by median (red line) with each dot representing one tumour and the number of tumours with signature indicated below. **c**, Profiles of the 8 DBS mutational signatures. D. Decomposed mutational signature landscape showing known and novel (top five) DBS signatures in relation to clinical and molecular parameters. Parameters shown by the proportion of tumours with the signature (circle size) and coloured by median tumour mutation burden (TMB).

---

**Supplementary Figure 4 (continued).** signatures from all tumours followed by estimation of the optimal solution (number of signatures marked with the grey line) based on the stability and accuracy of all 40 solutions. **b**, Mutation burden per megabase of each somatic SBS signature sorted by median (red line) with each dot representing one tumour and the number of tumours with signature indicated below. **c**, Profiles of the 27 SBS mutational signature. **d**, Decomposed mutational signature landscape showing known and novel (top two) SBS signatures in relation to clinical and molecular parameters. Parameters shown by proportion of tumours with the signature (circle size) and coloured by median tumour mutation burden (TMB).

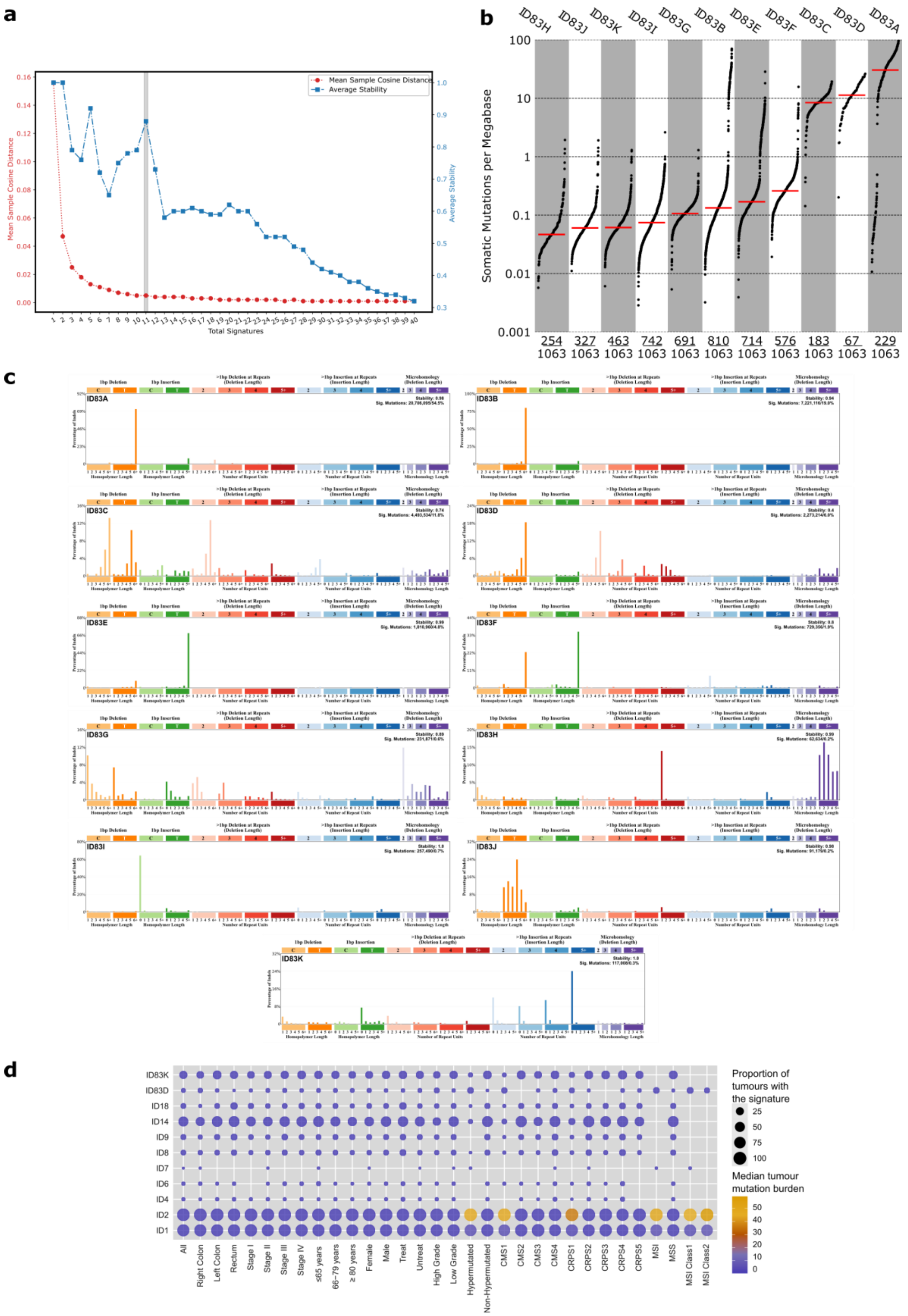


**Supplementary Figure 6.** ***De novo*** **extraction of small insertion and deletion (ID) mutational signatures**. **a**, Hierarchical *de novo* extraction of ID signatures from all samples followed by estimation of the optimal solution (number of signatures marked with the grey line) based on the stability and accuracy of all 40 solutions. **b**, Mutation burden per megabase of each somatic ID signature sorted by median (red line) with each dot representing one tumour and the number of tumours with signature indicated below. **c**, Profiles of the 11 ID mutational signatures. D. Decomposed mutational signature landscape showing known and novel (top two) ID signatures in relation to clinical and molecular parameters. Parameters shown by proportion of tumours with the signature (circle size) and coloured by median tumour mutation burden (TMB).

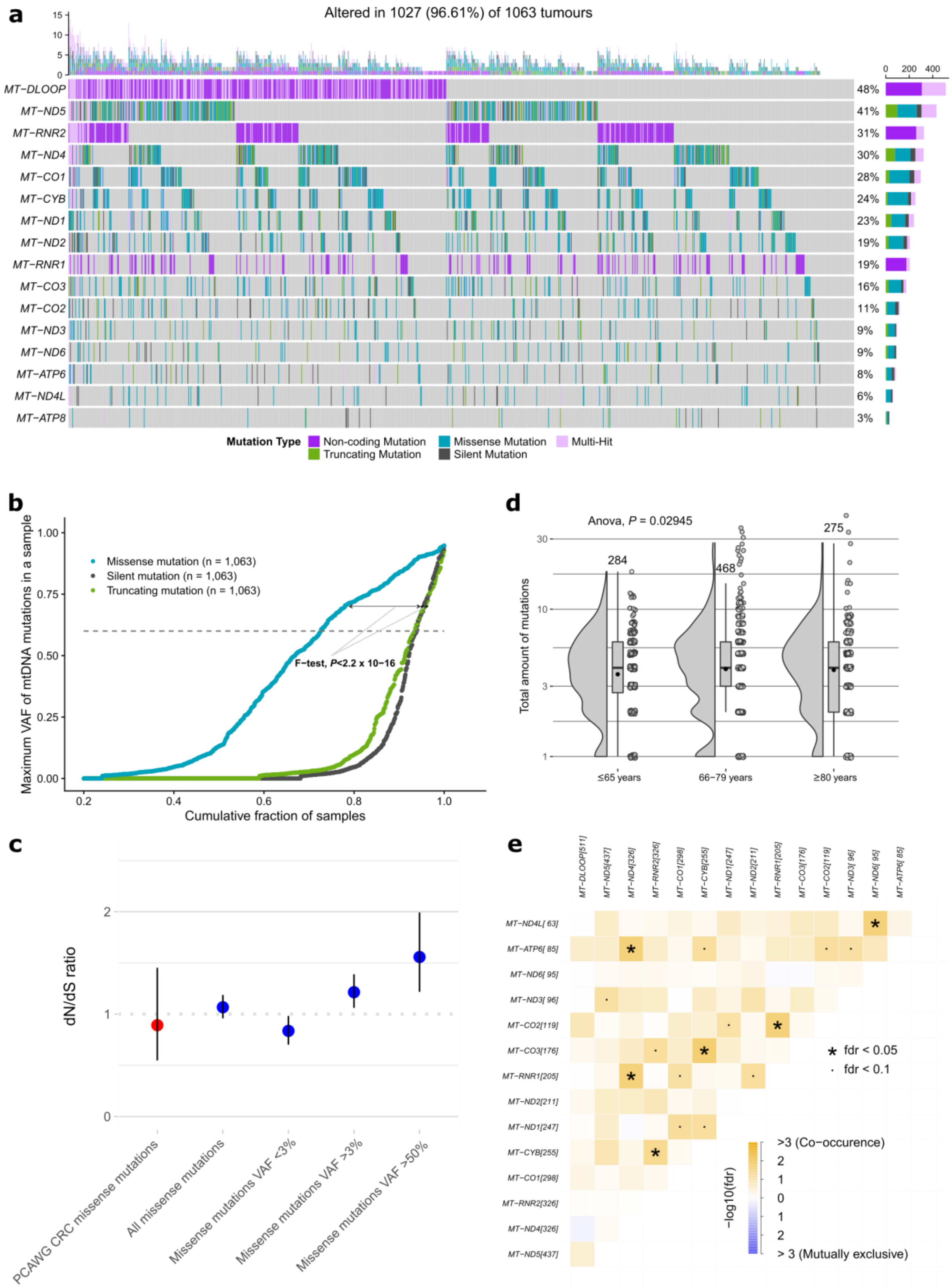


**Supplementary Figure 7. Somatic mutational landscape of mitochondrial genomes in colorectal cancer. a**, Oncoplot of somatic mitochondrial DNA gene (rows) mutations in 1,027 (97%) of the 1,063 sequenced tumours (columns). Tumour mutation burden for each tumour represented on the top and number of tumours with the mutation showed in the right, coloured by mutation type. **b**, Variant allele frequency (VAF) accumulation curves missense, silent and truncating mitochondria mutations (n, number of samples; F-test, Fisher's exact test). **c,** dN/dS ratio for mtDNA somatic missense mutations by different VAFs cut-offs. Error bars represent the 95% confidence intervals. **d,** Total amount of mitochondria mutations distribution per age group with multiple ANOVA comparison. The amount of tumours in each group is presented above the box plots and mean values as black dots. **e**, Mutually exclusive or co-occurring mitochondrial gene mutations in all tumours (* FDR<0.05 and ▪ FDR<0.1). The number of patients with the mutation is represented inside brackets next to the gene name.

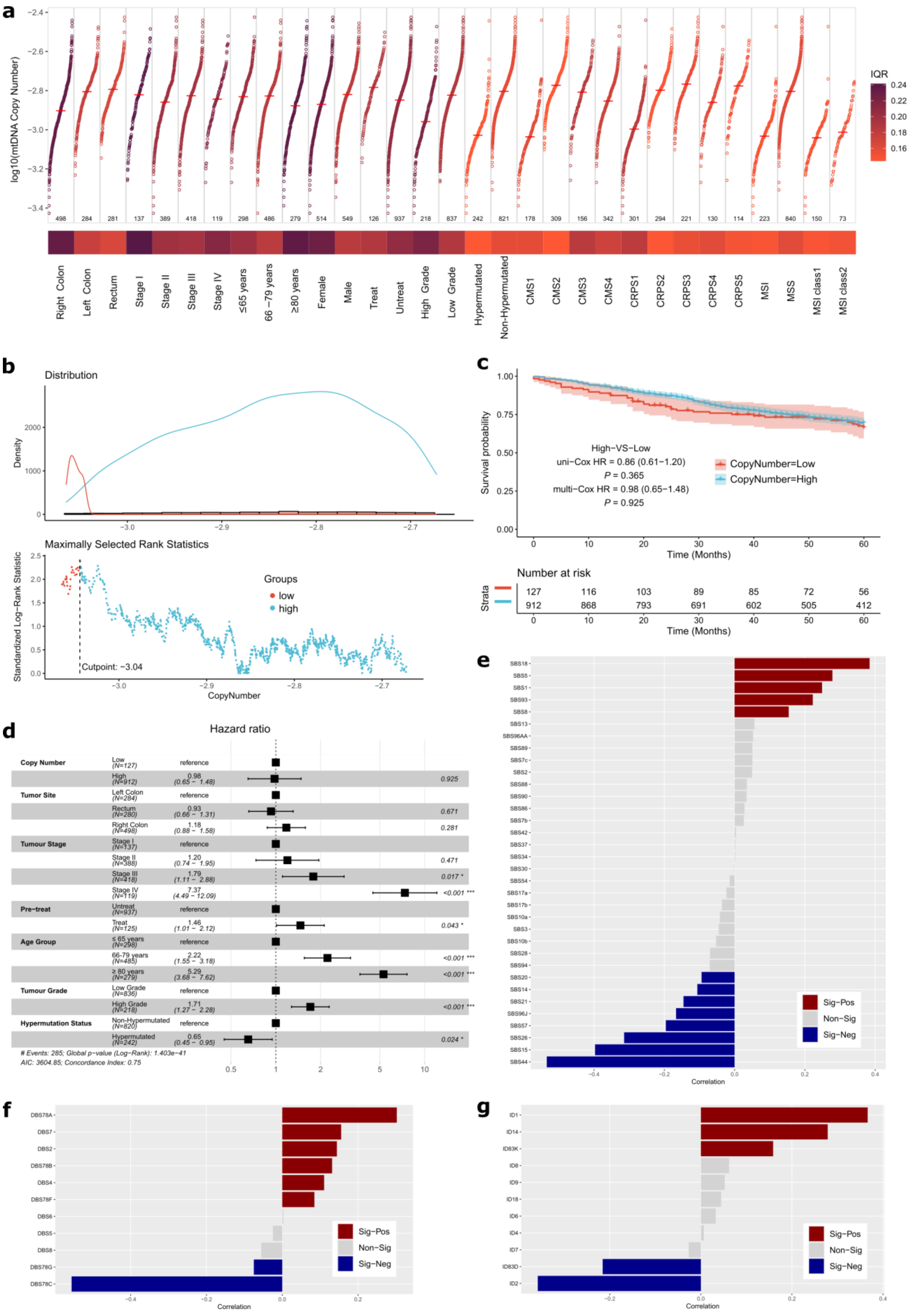


**Supplementary Figure 8. Mitochondrial copy number in 1,063 colorectal cancer tumours. a**, Mitochondrial genome copy number (mtDNA-CN)

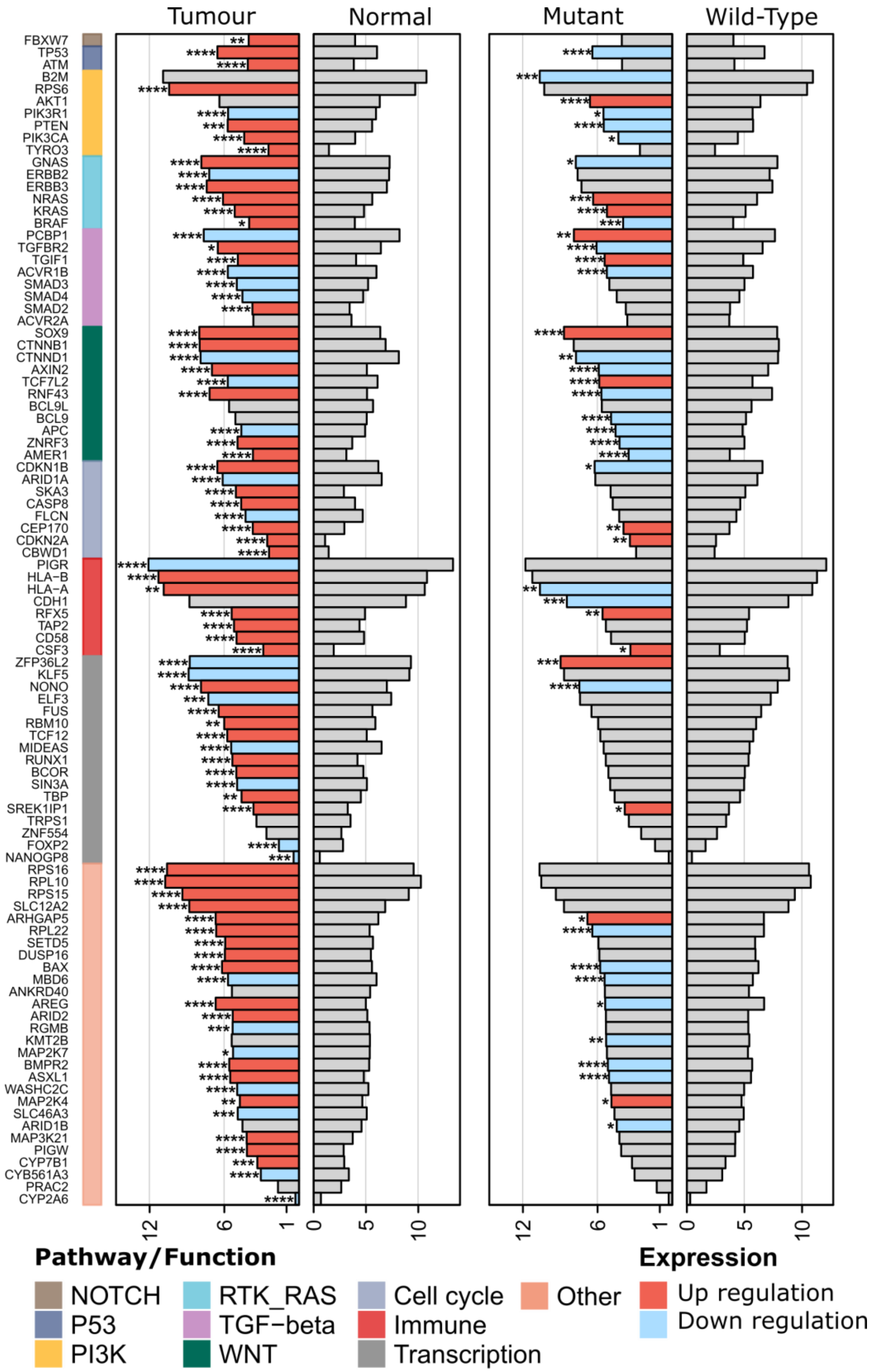

**Supplementary Figure 9. Gene expression profiles of the 96 driver genes.** Mean expression of driver genes in normal colorectal tissues (n=120) versus tumours (n=1,063) (left panel) and in wild-type (WT) versus mutant tumours (right panel). Genes were sorted by pathways/functions. Significance for differential gene expression was tested with Wilcoxon Rank Sum Test adjusted by Benjamini-Hochberg False Discovery Rate (* *P*<0.05, ** *P*<0.01, *** *P*<0.001, **** *P*<0.0001). Bars represented as log2(mean expression+1).

---

**Supplementary Figure 8 (continued).** landscape. For each feature, mtDNA-CN median was marked with a horizontal red line and variability was measured and coloured according to the interquartile range (IQR). Each dot represents a tumour and number included in each feature is shown in the bottom. **b**, A cut point (dashed line) was defined for low (red) and high (blue) mtDNA-CN according to their ranked density distribution. **c**, Overall survival for mtDNA-CN groups was calculated using Kaplan-Meier curves with respective confidence intervals (shading) and number of patients at

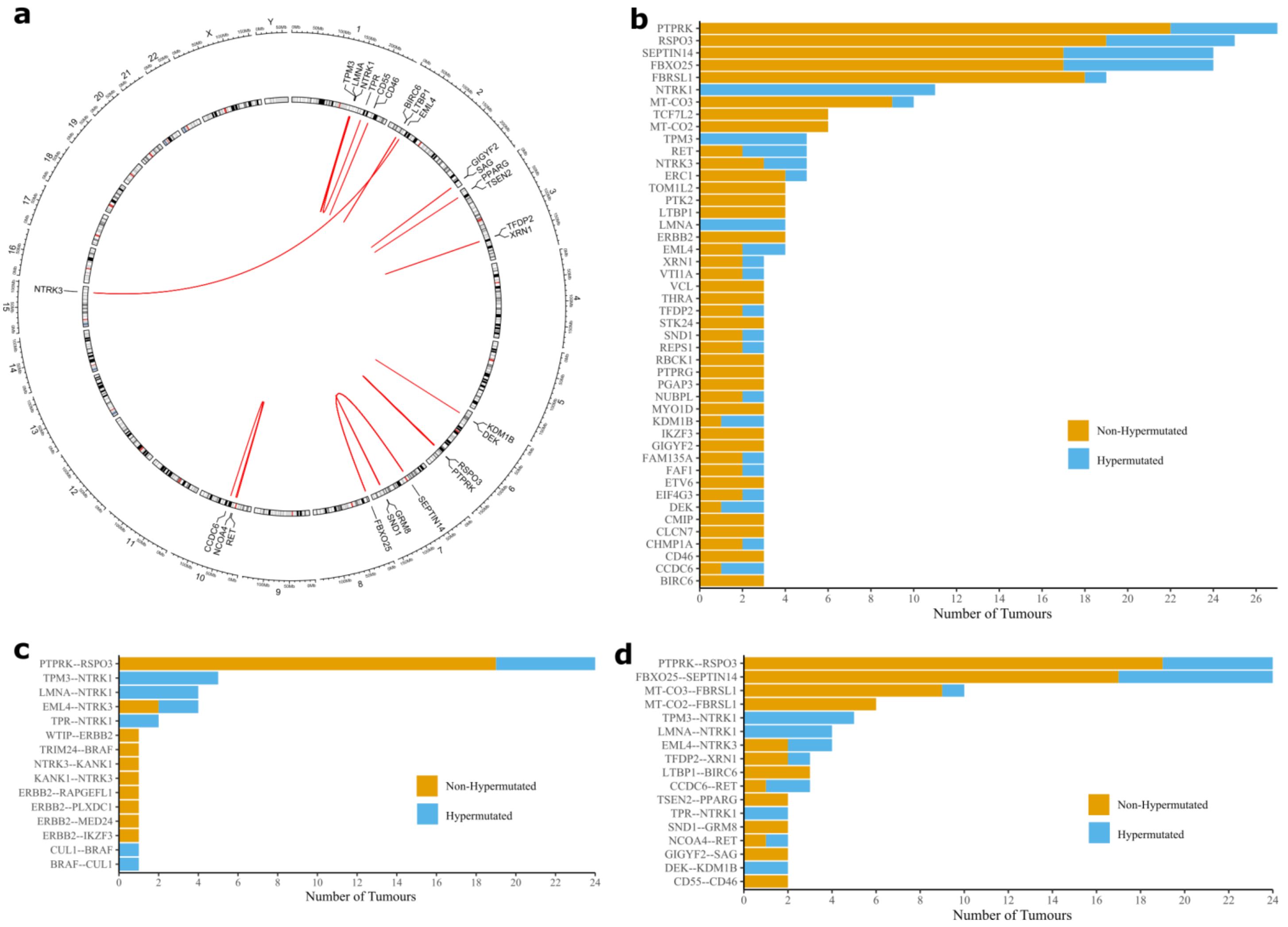


**Supplementary Figure 10. In-frame gene fusions detected through expression analysis.** Fusion genes were detected in RNAseq data from the 1,063 colorectal cancer tumours using STAR-Fusion and Arriba. **a**, Circos plot of recurrent fusions by chromosome location excluding mitochondrial DNA fusions. **b**, Top 50 genes recurrently included in fusions by hypermutation status. **c**, Recurrent colorectal cancer fusions described by Filippo *et al.* ([https://doi.org/10.3390/ijms20215319](https://doi.org/10.3390/ijms20215319)) observed in this cohort, with indication of hypermutation status. **d**, Top 20 recurrent expressed gene fusions, with indication of hypermutation status of affected samples.

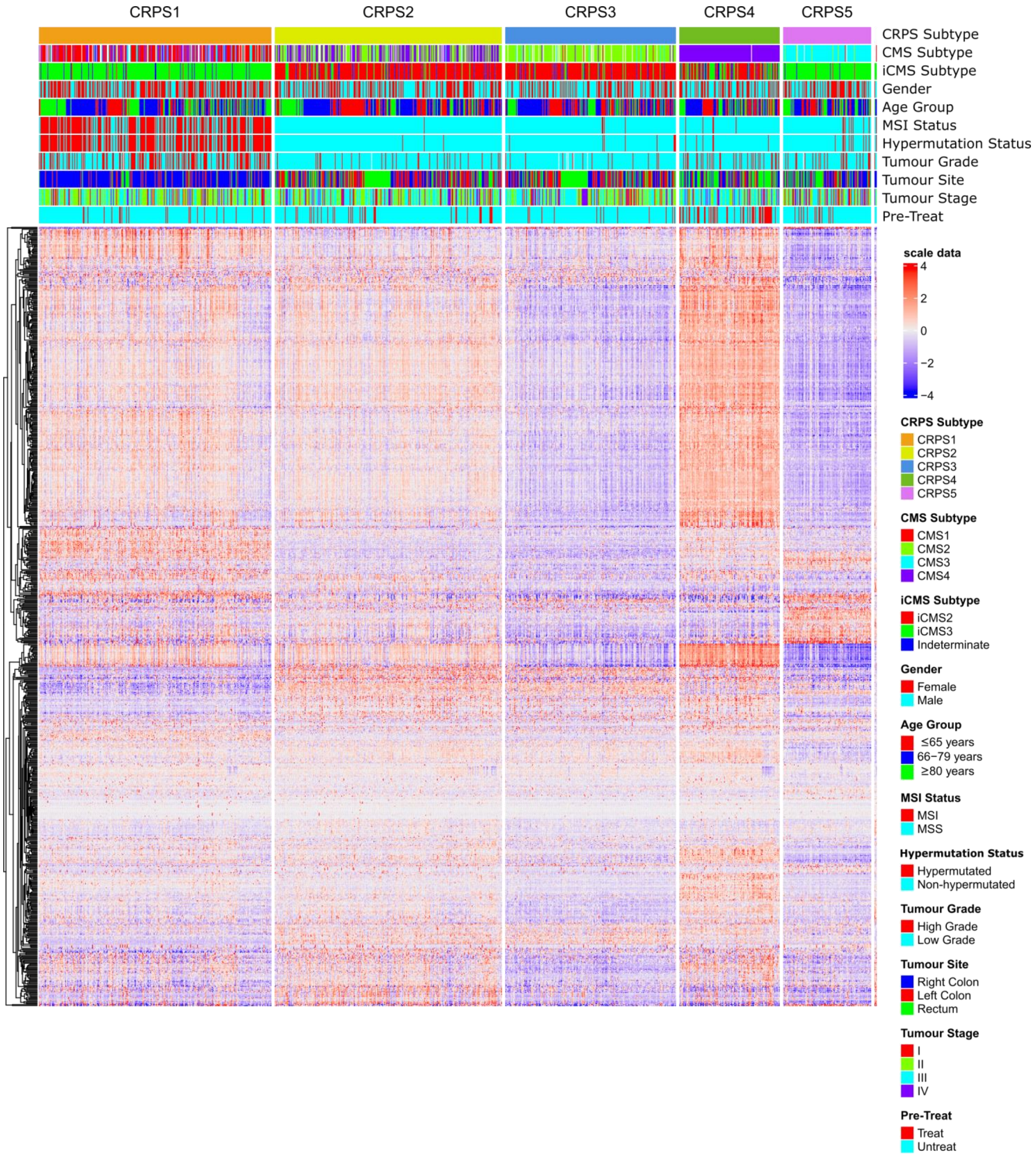


**Supplementary Figure 11. Colorectal cancer prognostic subtypes (CRPS).** Heatmap of the marker genes in each CRPS subtype for which red indicating high expression and blue low expression of a particular gene. Other clinical and genomic features represented above according to respective colour schemes. Subtypes were identified by unsupervised clustering of transcriptomes from 1,063 CRCs using Seurat (version 4.1.0). Potential batch effects or source differences between samples were corrected by Celligner. The stability of the clusters was assessed by Jaccard similarity index and the preferred clustering result (resolution=0.9, PC=20, K=20) was determined by scclusteval. The Intrinsic CMS (iCMS) subtype classification was performed as previously described (https://doi.org/10.1038/s41588-022-01100-4) and cases were defined as indeterminate if permutation-based FDR≥0.05.

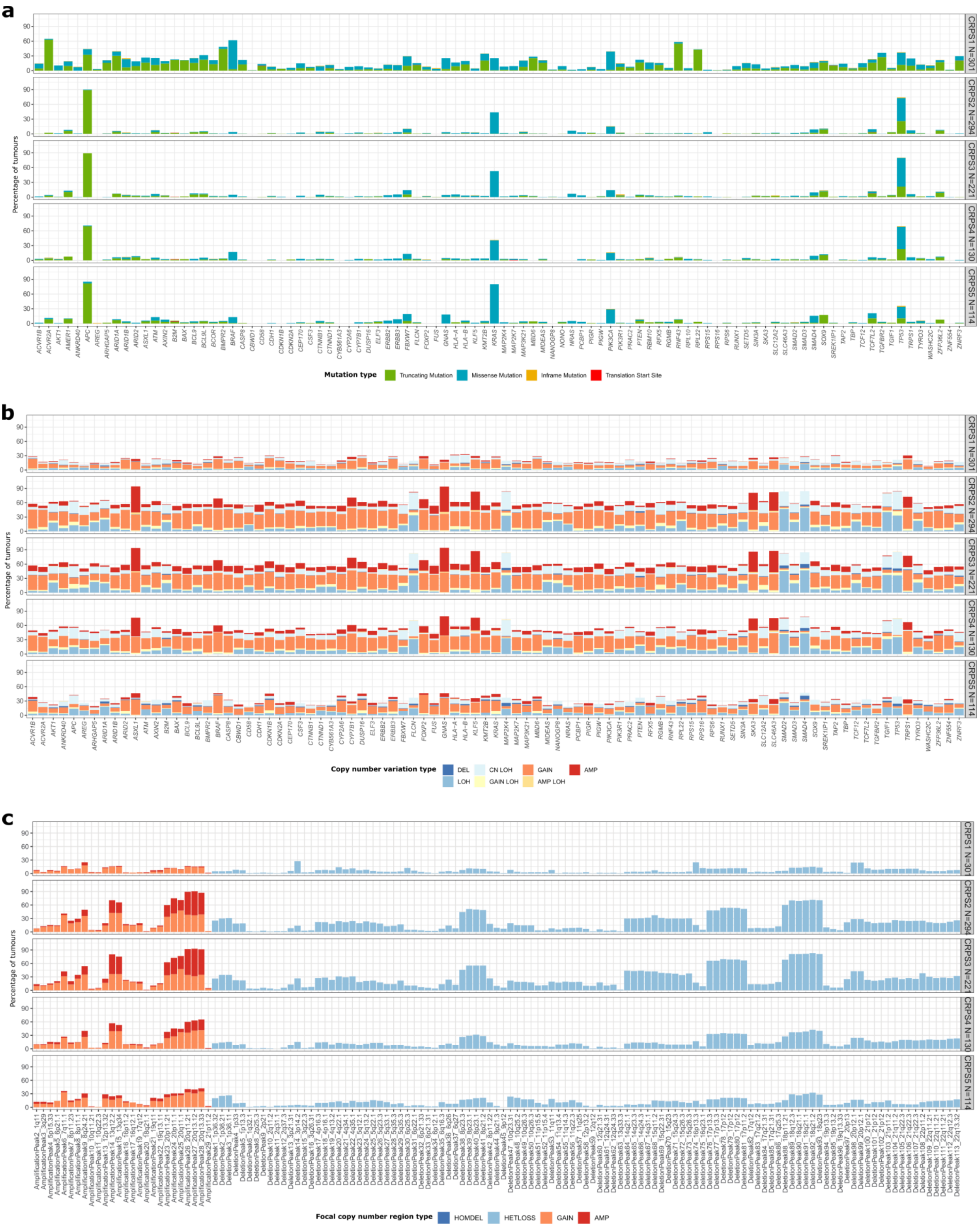


**Supplementary Figure 12. Somatic mutations and copy number variation by colorectal cancer prognostic subtypes (CRPS)**.

**Supplementary Figure 12 (continued). a**, Somatic mutations in 96 driver genes for the 1,063 colorectal cancer tumours by CRPS subtype. **b**, Frequency and type of somatic copy number variation in 96 driver genes by CRPS subtype. **c**, Focal copy number regions by CRPS subtype determined by GISTIC if Q<0.1. DEL, deletion; LOH, loss of heterozygosity; cn, copy number neutral; AMP, amplification; HOMDEL, homozygous deletion; HETLOSS, heterozygous deletion.

**Supplementary Figure 13 (continued)**. tumours, respectively were excluded. This leads to similar CRPS and CMS (head row) incidences as the original clustering where all cases, untreated and pre-treated at stages I-IV were included. **b**, In total, eleven colorectal cancer external datasets (n = 2,661 patients) from both NCBI GEO and NCI Genomic Data Commons were uniformly processed from FPKM and transformed to pathway profiles with ssGSEA. Comparison between CRPS and CMS classification for all external datasets (left) and the TCGA COAD/READ dataset only (right). The samples were coloured after their CMS subtype. **c,** Overall survival by CRPS for external datasets was calculated with Kaplan-Meier curves and multivariable log-rank test. D. Comparison of CMS Gene-Set activities using CMScaller (version v0.9.2) for the TCGA dataset (left) and this cohort (right) by CRPS subgroup (columns). Upregulation marked in red and downregulation as blue for each activity by row. NA, undefined subtype.

**a**

***Re-Cluster CRPS for Stage I - III Tumours Only***

| | CRPS1 | CRPS2 | CRPS3 | CRPS4 | CRPS5 | CRPSn | | CMS1 | CMS2 | CMS3 | CMS4 | NA |
|---|---|---|---|---|---|---|---|---|---|---|---|---|
| *ReClusters_CRPS1* | 245 | 13 | 0 | 3 | 1 | 0 | *ReClusters_CRPS1* | 140 | 1 | 11 | 44 | 66 |
| *ReClusters_CRPS2* | 3 | 240 | 9 | 0 | 0 | 0 | *ReClusters_CRPS2* | 0 | 110 | 1 | 82 | 59 |
| *ReClusters_CRPS3* | 1 | 5 | 185 | 0 | 2 | 0 | *ReClusters_CRPS3* | 2 | 140 | 15 | 6 | 30 |
| *ReClusters_CRPS4* | 0 | 4 | 0 | 107 | 0 | 0 | *ReClusters_CRPS4* | 0 | 0 | 0 | 107 | 4 |
| *ReClusters_CRPS5* | 27 | 1 | 1 | 0 | 100 | 0 | *ReClusters_CRPS5* | 2 | 2 | 111 | 0 | 14 |
| *ReClusters_CRPSn* | 0 | 1 | 0 | 1 | 1 | 119 | *ReClusters_CRPSn* | 1 | 0 | 44 | 16 | 61 |

***Re-Cluster CRPS for Untreat Tumours Only***

| | CRPS1 | CRPS2 | CRPS3 | CRPS4 | CRPS5 | CRPSn | | CMS1 | CMS2 | CMS3 | CMS4 | NA |
|---|---|---|---|---|---|---|---|---|---|---|---|---|
| *ReClusters_CRPS1* | 235 | 1 | 0 | 5 | 0 | 0 | *ReClusters_CRPS1* | 138 | 0 | 24 | 30 | 49 |
| *ReClusters_CRPS2* | 3 | 209 | 4 | 11 | 3 | 0 | *ReClusters_CRPS2* | 0 | 99 | 5 | 76 | 50 |
| *ReClusters_CRPS3* | 1 | 6 | 177 | 0 | 4 | 0 | *ReClusters_CRPS3* | 3 | 136 | 15 | 7 | 27 |
| *ReClusters_CRPS4* | 37 | 33 | 0 | 66 | 1 | 0 | *ReClusters_CRPS4* | 1 | 1 | 3 | 101 | 31 |
| *ReClusters_CRPS5* | 1 | 0 | 0 | 0 | 88 | 0 | *ReClusters_CRPS5* | 0 | 1 | 83 | 0 | 5 |
| *ReClusters_CRPSn* | 0 | 0 | 0 | 1 | 1 | 120 | *ReClusters_CRPSn* | 1 | 0 | 43 | 16 | 62 |

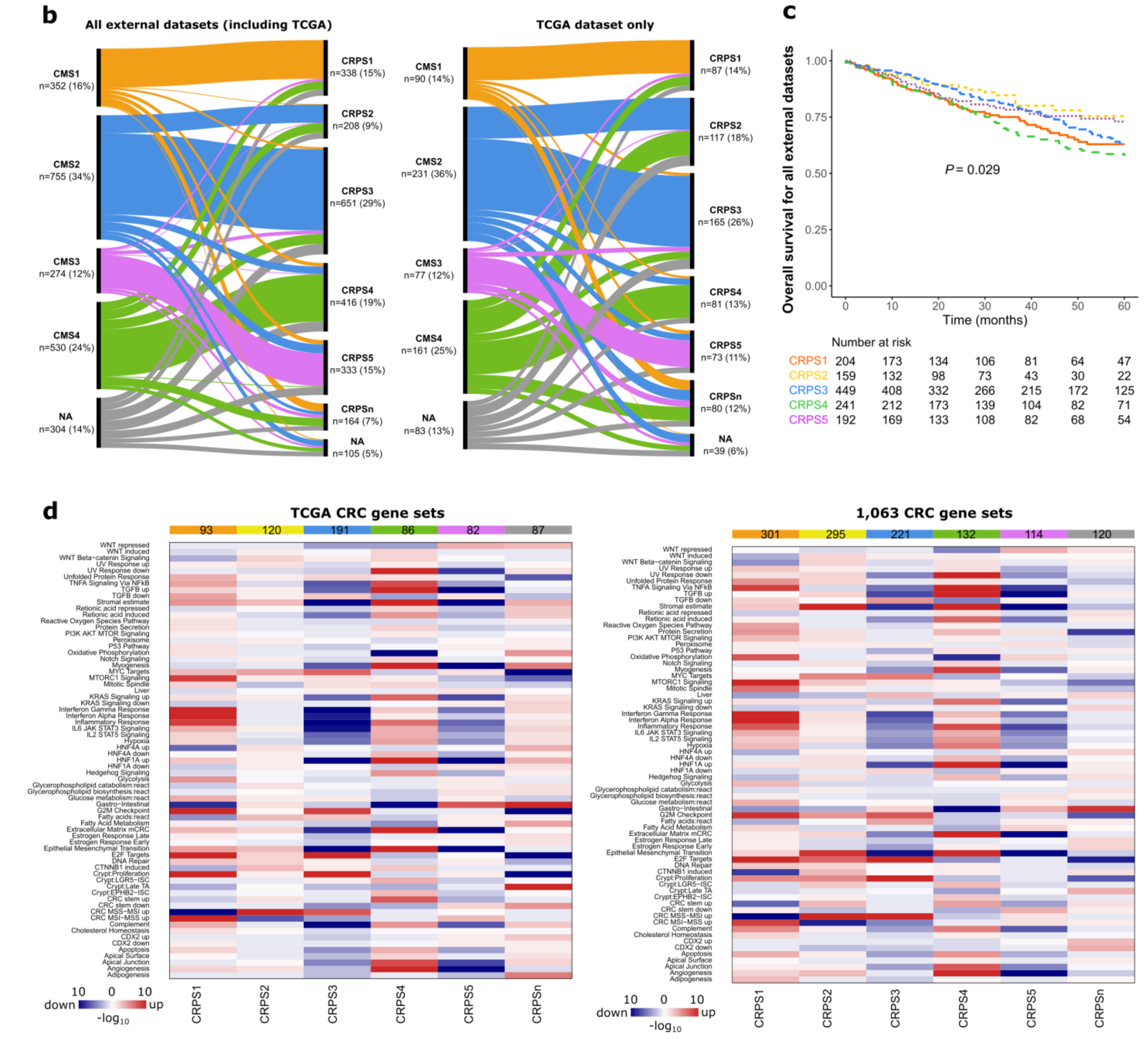


**Supplementary Figure 13. Validation of CRPS**. **a**, Distribution of cases from re-clustering by CRPS (first column) when stage IV and pre-treated

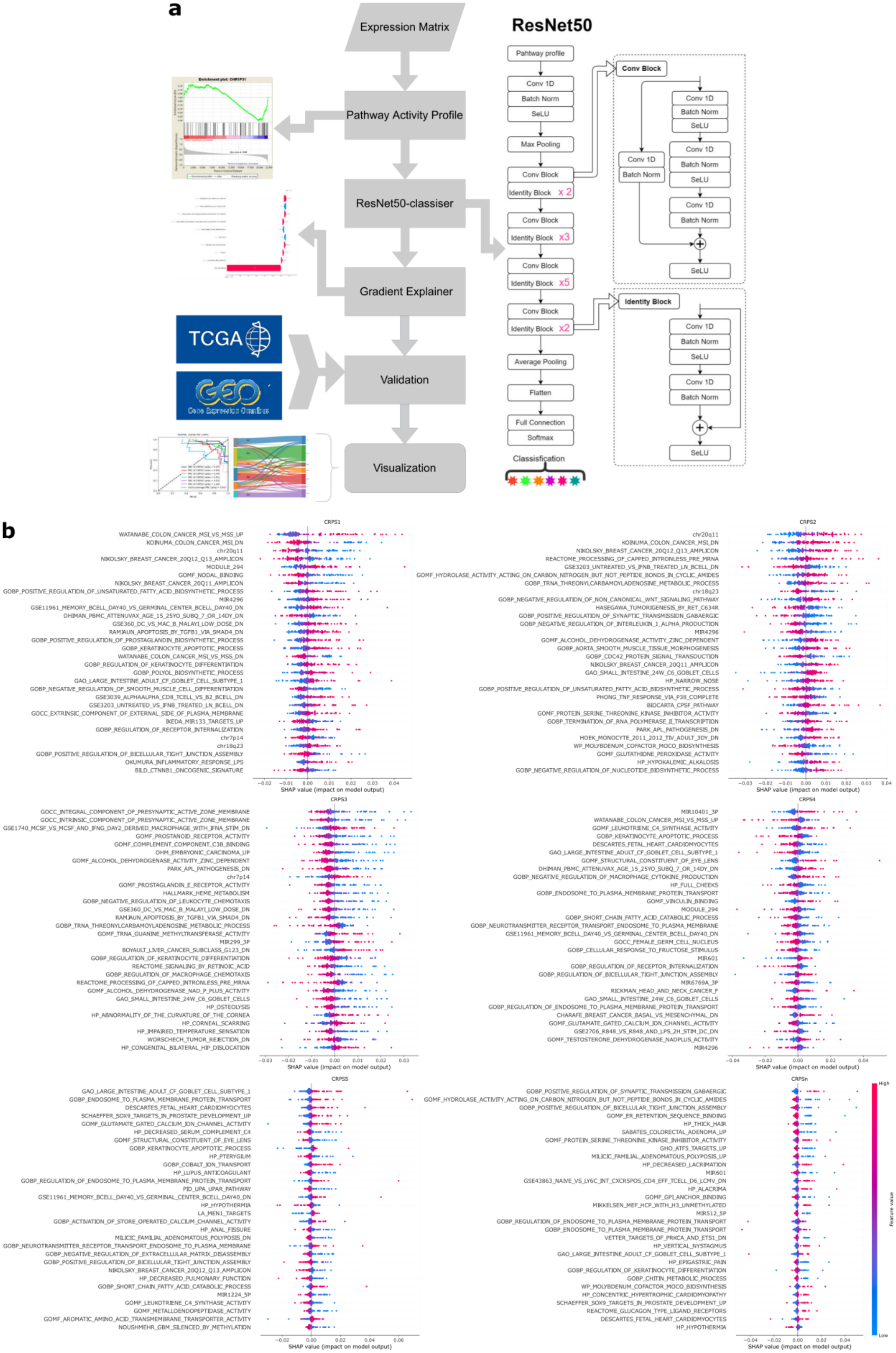


**Supplementary Figure 14. Deep learning model for CRPS classification and model interpretation at the pathway level. a**, A deep neural network

**Supplementary Figure 14 (continued).** architecture for the CRPS classifier was built with gene expression data from the 1,063 colorectal cancers based on the ResNet50 deep residual learning framework. For more detailed step information see Methods. **b**, Shapley Additive exPlanations (SHAP) was applied to CRPS classifications to explain model predictions. SHAP values are shown for the features that contributed significantly for each CRPS subtype and coloured by feature value. CRPSn, CRPS normal sample subtype.

**Supplementary Figure 15 (continued).** mRNA abundance signature (see Methods) were calculated for all tumours (top) and correlated with (**a**) mutations in the 96 driver genes, (**b**) mutation burden and structural variants, and (**c**) numbers of mutations attributed as clonal and subclonal. Adjusted FDR *P*-values shown to the right and significance threshold indicated by dotted line. PGA, percentage of genome with copy number alterations; CNA, copy number alterations; SNV, single nucleotide mutation; DNV, double nucleotide mutation; TNV, triple nucleotide mutation; DEL, deletion; INS, insertion; INDEL, insertion and deletion.

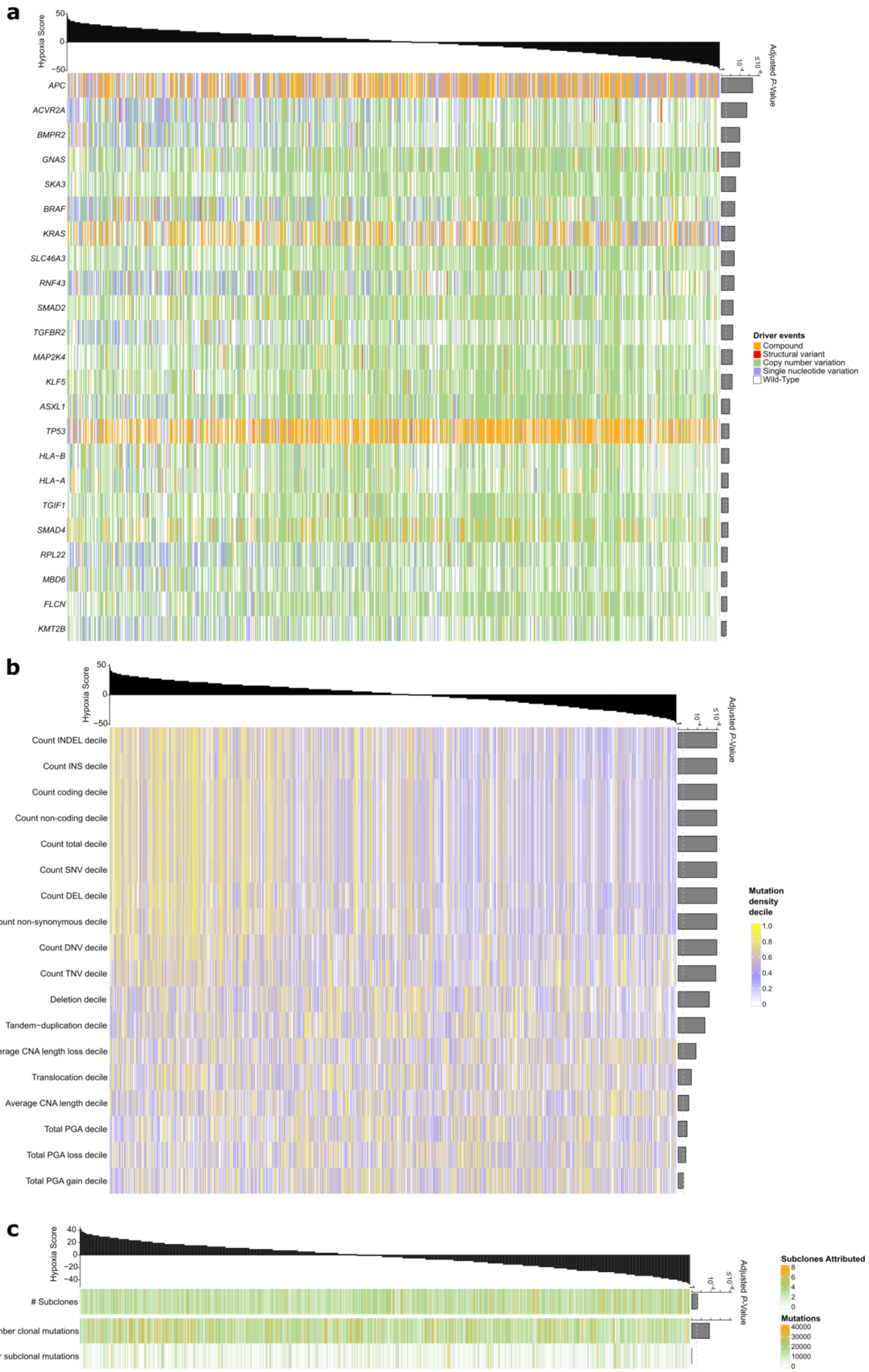


**Supplementary Figure 15. Hypoxia correlation with mutations, structural variants and mutational clonality.** Hypoxia scores based on the Buffa

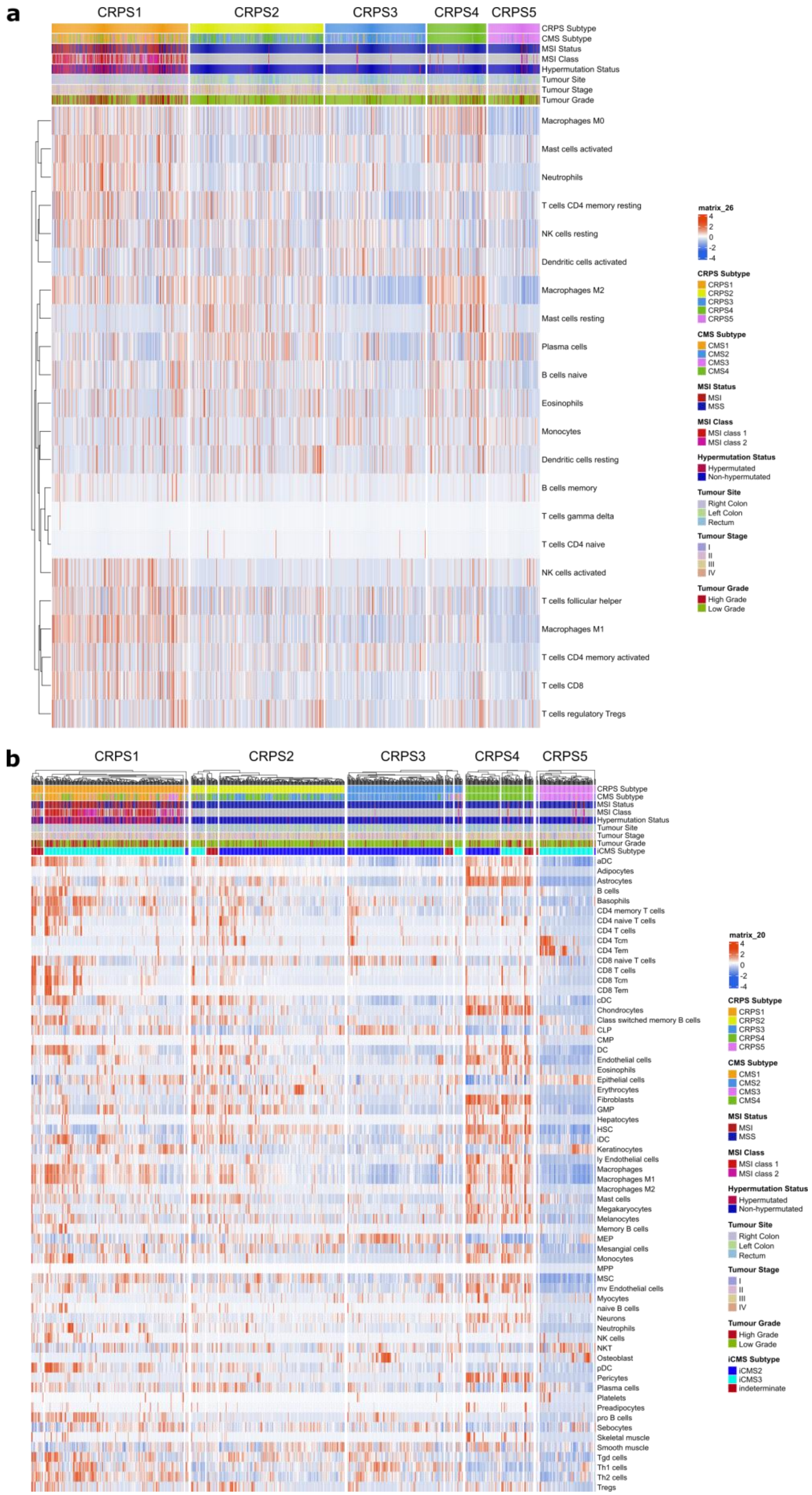


**Supplementary Figure 16. Immune and stromal cell composition by CRPS**. Immune and stromal cells were predicted for each CRPS subtype using

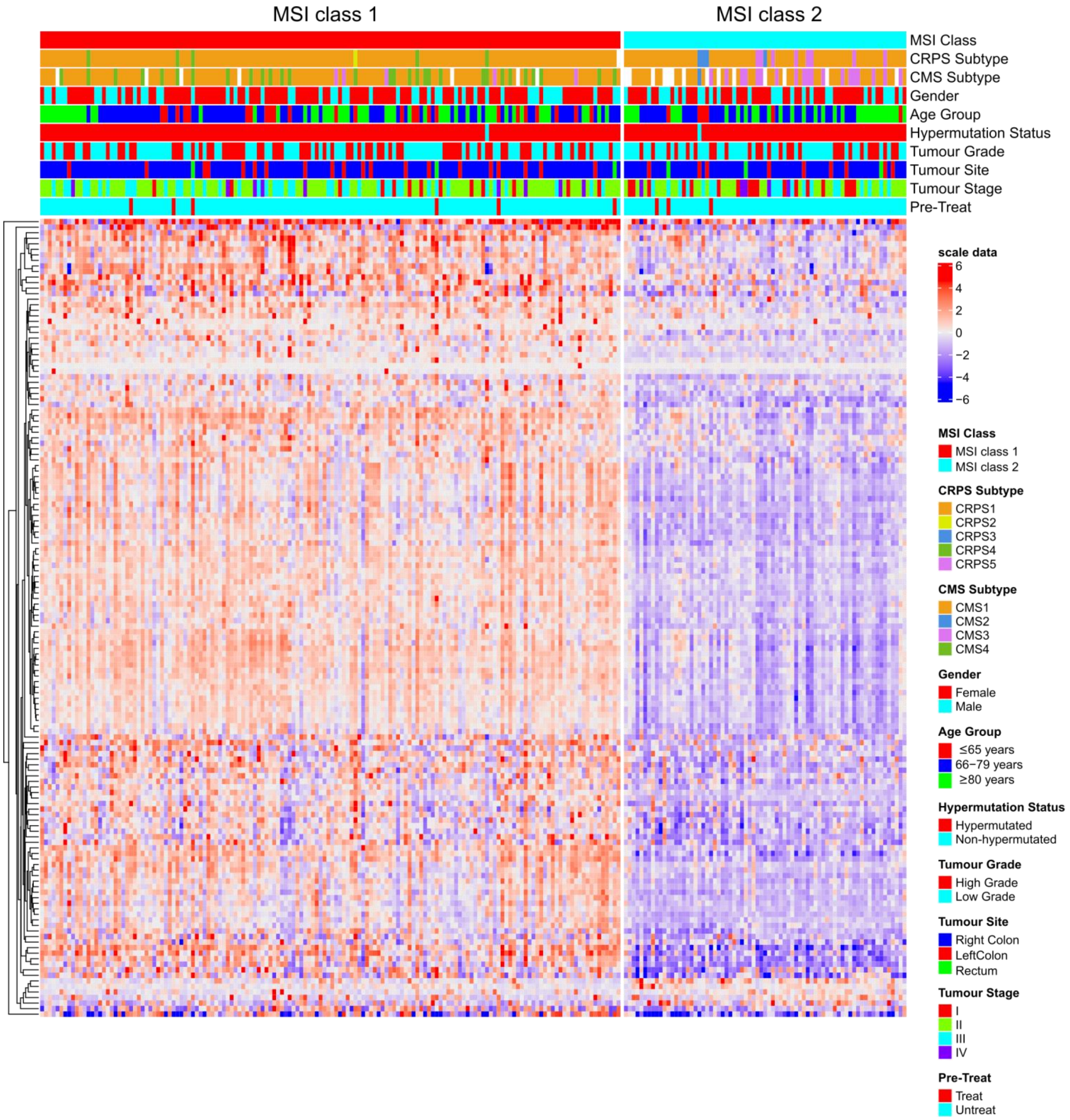


**Supplementary Figure 17. Microsatellite instable colorectal cancers clustering classification.** Unsupervised clustering of transcriptome data divided microsatellite instable (MSI) colorectal cancer samples in two classes defined as class 1 (left) and class 2 (right). Heatmap of the top 150 marker genes in each MSI class for which red indicating high expression and blue low expression of a particular gene. Other clinical and genomic features represented above according to respective colour schemes.

---

**Supplementary Figure 16 (continued).** gene expression data and the (**a**) CIBERSORT and (**b**) xCell algorithms. Each row represents a cell and tumours were grouped by CRPS subtype. Clinical and genomic features represented above each heatmap according to respective colour schemes. Intrinsic subtypes of CMS (iCMS) are also indicated above the heatmap in **b**.

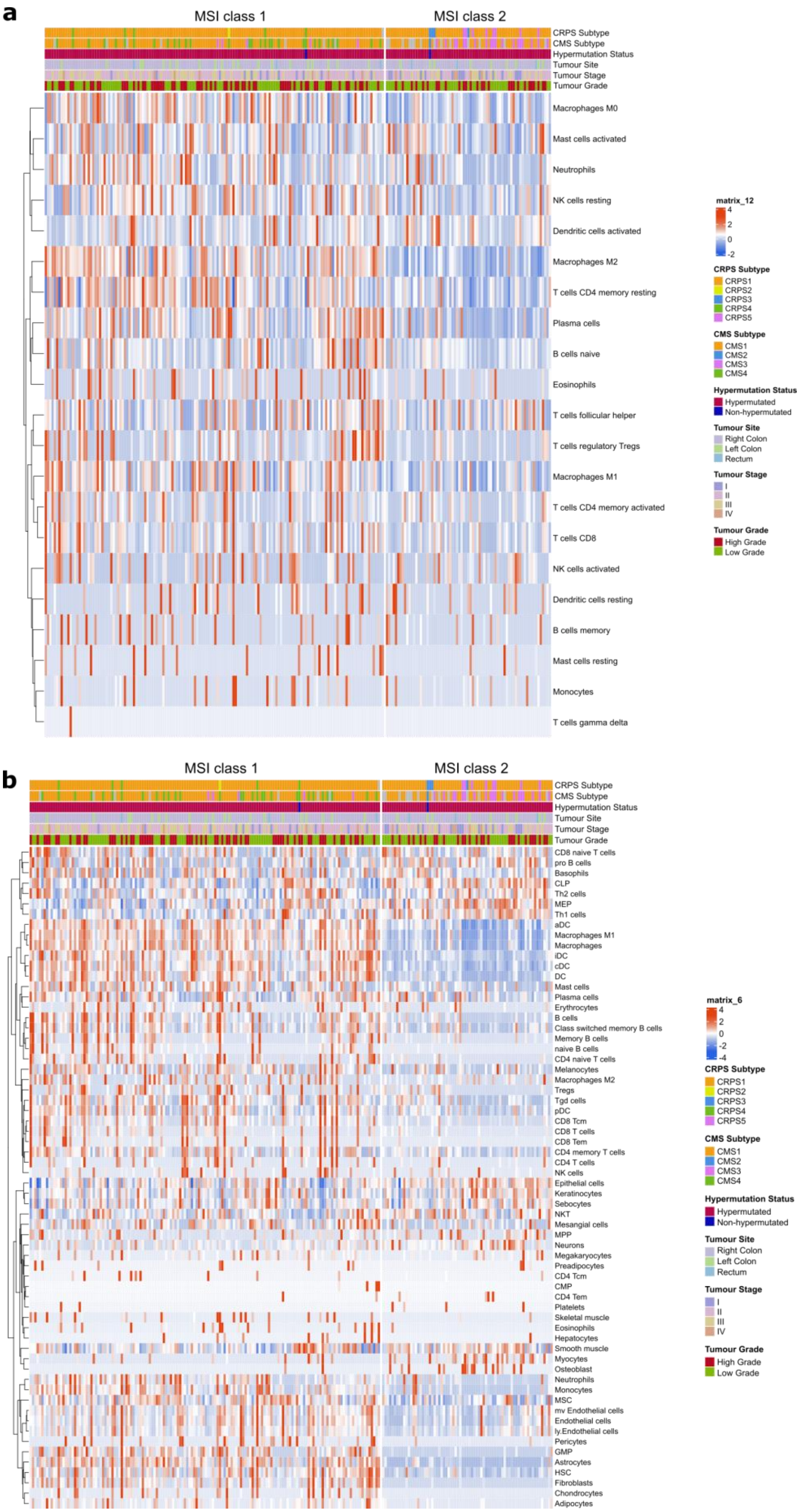


**Supplementary Figure 18. Infiltration of immune and stromal cell populations by microsatellite instability class**. Heatmaps of predictions of

**Supplementary Figure 18 (continued).** immune and stromal cell population for the two microsatellite instable (MSI) classes for (**a**) CIBERSORT and (**b**) xCell algorithms. Each row represents a cell and tumours were grouped by MSI class. Clinical and genomic features represented above each heatmap according to respective colour schemes. MSI class 1 samples had higher frequency of infiltrated lymphocytes and stromal cells compared to MSI class 2 samples.

**Supplementary Figure 19 (continued).** free survival by mismatch repair status and MSI class for cells predicted by CIBERSORT (left) and xCell (right) algorithms. Univariable Cox regression was performed on cell types that showed expression in at least 5 patients with survival data, and statistically significant differences (* $P$<0.05 and ** $P$<0.01) were further tested by multivariable Cox regression with co-variates including tumour site, treatment status, tumour stage, age groups, and tumour grade. The hazard ratio values are indicated by colour intensity. **b**, Percentage of tumours with somatic mutations in 96 driver genes for MSI class 1 (top) and class 2 (bottom) cases. **c**, Percentage of tumours with somatic copy number variation of 96 driver genes for MSI class 1 (top) and class 2 (bottom) cases. D. Percentage of tumour with focal copy number regions (Q<0.1) gained or lost, determined by GISTIC in the MSI class 1 (top) and class 2 (bottom) cases. LOH, loss of heterozygosity; cn, copy number neutral; AMP, amplification; HOMDEL, homozygous deletion; HETLOSS, heterozygous deletion.

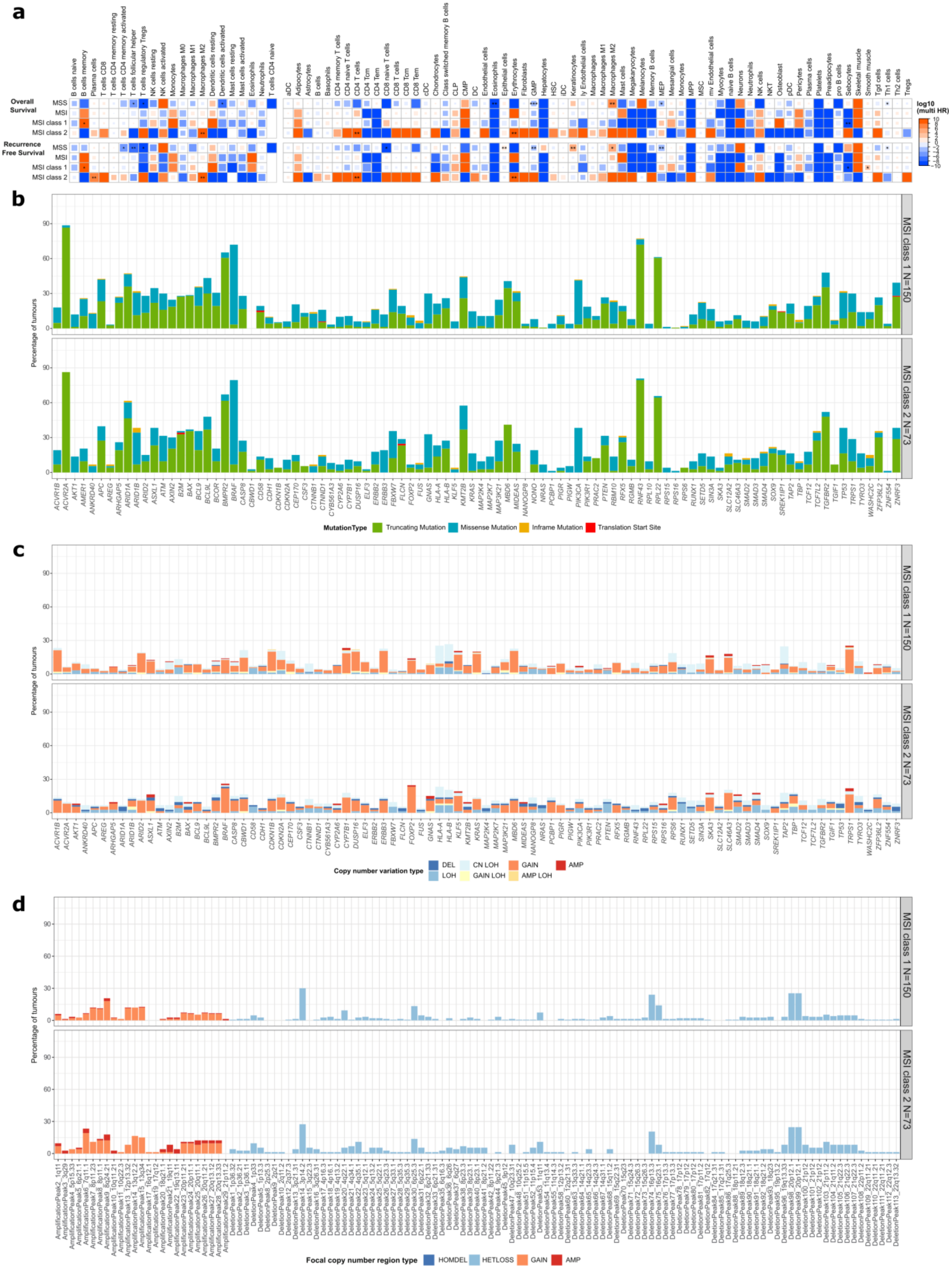


**Supplementary Figure 19. Survival, somatic mutations and copy number variation by microsatellite instability class**. **a**, Overall and recurrence
