## Supplementary Tables for "Prognostic whole-genome and transcriptome signatures in colorectal cancers": Supplementary_Table_13.docx

| **Supplementary Table 13. Cohort characteristics compared to surgically treat colorectal cancer patients in Sweden 2007-2016.** | | | |
| --- | --- | --- | --- |
| ***Characteristics*** | **Present Cohort**  **n =1,063** | **Sweden Resected Patients**  **2007-2016**  **n = 48,470** | ***P*-value** |
| ***Age (years)*** |  |  |  |
| *>75 years* | 422 (40) | 18,751 (39) | 0.504 |
| ***Sex*** |  |  |  |
| *Female* | 514 (48) | 23,091 (48) | 0.664 |
| *Male* | 549 (52) | 25,379 (52) |  |
| ***Primary Tumour Location*** |  |  |  |
| *Colon* | 782 (74) | 33,957 (70) | **0.01** |
| *Rectum* | 281 (26) | 14,513 (30) |  |
| ***Tumour Stage*** |  |  |  |
| *Stage I* | 138 (13) | 9,103 (19) | **1.9e-7** |
| *Stage II* | 392 (37) | 16,404 (34) |  |
| *Stage III* | 419 (39) | 16,742 (34) |  |
| *Stage IV* | 114 (11) | 6,221 (13) |  |
| ***Surgery*** |  |  |  |
| *Tumour surgically removed* | 1,034 (97) | 48,470 (100) | - |
| *Not operated* | 29 (3) | - |  |
| ***5-year overall survival (OS)*** |  |  |  |
| *All Patients %* | 69% | 67% | - |
| *Stage I-III %* | 75% | 72% | - |
| *Stage IV %* | 24% | 28% | - |

Data from the Swedish Regional Cancer Center (RCC) quality register interactive report of the Swedish Colorectal Cancer Register (SCRCR) for colon and rectal cancers.
