## Supplementary Tables for "Prognostic whole-genome and transcriptome signatures in colorectal cancers": Supplementary_Table_14.docx

| **Supplementary Table 14.** Clinical characteristics of 114 colorectal cancer patients with synchronous metastases. | | | | |
| --- | --- | --- | --- | --- |
| ***Characteristics*** | **Right Colon**  **n = 43 (38%)** | **Left Colon**  **n = 36 (31%)** | **Rectum**  **n = 35 (31%)** | ***p*-value** |
| ***Age (years)*** |  |  |  |  |
| *Mean ± S.D.* | 70.6 ± 11.5 | 67.6 ± 10.7 | 71.9 ± 8.5 |  |
| *Median (Range)* | 70 (44-89) | 68.5 (42-86) | 73 (56-88) |  |
| *>75 years* | 17 (40) | 9 (25) | 12 (34) | 0.405 |
| ***Sex*** |  |  |  |  |
| *Female* | 27 (63) | 17 (47) | 14 (40) | 0.114 |
| ***Histology Subtype*** |  |  |  |  |
| *Adenocarcinoma* | 30 (70) | 29 (81) | 32 (91) | 0.060 |
| *Mucinous adenocarcinoma* | 13 (30) | 7 (19) | 3 (9) |  |
| ***Tumour Grade*** |  |  |  |  |
| *Low grade malignancy* | 20 (47) | 30 (83) | 28 (80) | **0.0005** |
| ***Pre-Treated*** |  |  |  |  |
| *Yes* | 7 (16) | 7 (19) | 2 (6) | 0.191 |
| ***Metastases Location*** |  |  |  |  |
| *Liver* | 33 (77) | 27 (75) | 30 (86) | 0.524 |
| *Lung* | 10 (23) | 11 (31) | 19 (54) | **0.016** |
| *Peritoneum* | 13 (30) | 8 (22) | 3 (9) | 0.055 |
| *Lymph Node* | 5 (12) | 3 (8) | 7 (20) | 0.345 |
| *Bone* | 1 (2) | 1 (3) | 3 (9) | 0.444 |
| *Other* | 5 (12) | 1 (3) | 1 (3) | 0.221 |
| ***Treatment*** |  |  |  |  |
| *Best Supportive Care (BSC)* | 11 (26) | 5 (14) | 9 (26) | 0.381 |
| *Metastasectomy* | 21 (49) | 18 (50) | 10 (29) | 0.119 |
| *1^st^ Line chemotherapy* | 21 (49) | 29 (81) | 22 (63) | **0.014** |
| *2^nd^ Line* | 15 (35) | 14 (39) | 13 (37) | 0.969 |
| *3^rd^ Line ^+^* | 10 (23) | 6 (17) | 4 (11) | 0.414 |
| ***Median overall survival*** *(95% CI)* |  |  |  |  |
| *All* | 20 m (15-36) | 32 m (27-58) | 28 m (16 -31) | 0.224 |
| *BSC only* | 10 m (1-18) | 21 m (7-41) | 10 m (7-34) | 0.239 |
| *Treated with metastasectomy* | 55 m (28-n/a) | 58 m (30-n/a) | 64 m (21-n/a) | 0.885 |
| *Treated without metastasectomy* | 16 m (7-21) | 31 m (20-n/a) | 28 m (11-30) | 0.068 |

* BSC means that the patient did not have metastatic surgery or received tumour controlling systemic chemotherapy, whereas palliative radiotherapy could be provided. Most patients having metastasectomy also received peri-operative chemotherapy.
